# AGREEMENT AND ERROR RATES IN ANTIMICROBIAL SUSCEPTIBILITY TESTING FOR THREE COMMERCIAL AUTOMATED SYSTEMS: A SYSTEMATIC LITERATURE REVIEW AND META-ANALYSIS

**DOI:** 10.64898/2026.03.24.26349209

**Authors:** Kayla Van Benten, Lauren Cooper, Kayla Kirby, Shelby Kruer, Karen Byron

**Affiliations:** Waters Advanced Diagnostics (formerly BD Diagnostic Solutions), Sparks, MD, USA; Becton, Dickinson and Company, Franklin Lakes, NJ, USA

**Author notes:** **CORRESPONDENCE:** Kayla Van Benten, Waters Advanced Diagnostics (formerly BD Diagnostic Solutions), Sparks, MD 21152, USA.

**Keywords:** Antimicrobial susceptibility testing Clinical microbiology, Diagnostic agreement, Diagnostic error, Drug resistant bacteria, Automated antimicrobial susceptibility testing

## Abstract

**BACKGROUND:** Automated antimicrobial susceptibility testing (AST) systems are crucial for accurate, timely detection of drug-resistant microbial isolates. This meta-analysis assessed the performance of the BD Phoenix™ (“Phoenix”, BD Diagnostic Solutions), Vitek^®^ 2 (“Vitek 2”, bioMérieux), and DxM MicroScan WalkAway (“MicroScan”, Beckman Coulter, Inc.) AST systems relative to common reference methodology.

**METHODS:** A systematic literature search in Ovid (MEDLINE and Embase) yielded 275 unique (not duplicated) records, with 44 additional records retrieved from handsearching; 39 studies met inclusion criteria. Categorical agreement (CA), essential agreement (EA), very major errors (VMEs), and major errors (MEs) for the three instruments were compared to a common reference method. Ratios of proportions were analyzed using random-effect meta-regression.

**RESULTS:** The instruments did not differ significantly in CA, EA, or ME. Vitek 2 showed a higher overall VME rate than Phoenix (∼44% higher; Vitek 2-to-Phoenix ratio = 1.44; *p*=0.062 [approaching significance]) and MicroScan (74% higher; ratio = 1.74; *p*=0.045). No appreciable difference was observed for VME between Phoenix and MicroScan. Subgroup analyses should be interpreted cautiously due to limited overall significance indicating varying performance across systems. Vitek 2 generally had higher relative VMEs for gram-negative organisms and lower relative VMEs for gram-positive organisms, whereas Phoenix showed the opposite pattern. MicroScan had relatively low VMEs when stratified by Clinical and Laboratory Standards Institute (CLSI) criteria; no differences in VMEs were observed using European Committee on Antimicrobial Susceptibility Testing (EUCAST) criteria.

**CONCLUSION:** Although some VME differences were noted, overall performance of the three systems was comparable. Organism- and drug-specific VME patterns—and updates to CLSI criteria over time—highlight the importance of continued monitoring of current breakpoints for all three instruments.

## INTRODUCTION

Antimicrobial resistance (AMR) is a major public health challenge that threatens the efficacy of antimicrobial therapies. Currently, in the United States, there are over 2.8 million antimicrobial-resistant infections, leading to over 35,000 deaths annually.^1–3^ Globally, over 700,000 people die each year from drug-resistant diseases.^3^ These numbers highlight the urgent need for a worldwide response to AMR. Moreover, improper use and overuse of antibiotics can result in the emergence of drug-resistant pathogens,^1–3^ and lead to prolonged hospital stays, rising medical expenses, and increased mortality. As a result, infections that previously required routine treatment are becoming difficult to address.^3^ AMR also threatens the success of major surgical and cancer interventions that depend on effective antibiotics to prevent and treat infection.^1–4^ Therefore, combating AMR will require a global and coordinated response to ensure prudent use of antimicrobials, improve infection prevention and control, and speed up investment in new treatments.^1–3^ As part of this effort, clinical microbiology laboratories play a critical role in guiding appropriate therapy through timely and accurate identification and antimicrobial susceptibility testing (AST).^5^ At the same time, these laboratories are expected to operate with greater efficiency—delivering results more quickly, with fewer errors, and under increasing resource constraints.^6,7^ To meet these challenges, automated and semi-automated systems have been developed and implemented to improve accuracy and efficiency of pathogen detection while streamlining clinical workflow and reducing the time required for sample preparation and processing.^8^ Although automated AST systems have improved laboratory operations, the persistent evolution of bacterial resistance presents a shared challenge for laboratories and manufacturers to ensure these systems remain clinically relevant and responsive to emerging threats.

This systematic literature review (SLR) and meta-analysis was performed to compare the performance of three commercial AST instruments: Vitek^®^ 2 (“Vitek 2;” bioMérieux),^9^ BD Phoenix™ (“Phoenix;” Becton, Dickinson and Company),^10^ and DxM MicroScan WalkAway (“MicroScan;” Beckman Coulter)^11^. A systematic literature search was conducted for articles reporting on at least two of the above-mentioned instruments, and data pertaining to categorical agreement (CA), essential agreement (EA), very major errors (VMEs), and major errors (MEs) were captured and compared.

## MATERIALS AND METHODS

This study followed the standards outlined by the Cochrane Collaboration Diagnostic Test Accuracy Working Group for the conduct of meta-analysis as well as the PRISMA guidelines for reporting SLR results.^12–14^ The protocol for this systematic literature review was registered with PROSPERO International Prospective Register of Systematic Reviews (PROSPERO CRD4202460315).^15^

The PICO (Participants; Interventions; Comparators; Outcomes) for this SLR was developed during conception of the protocol: *Participants* were those providing a specimen that showed a positive result for infection with identification of the infectious microorganisms; *Intervention* was AST associated with the positive sample to determine appropriate therapy by identifying effective agents and resistance profiles, with three common commercial platforms (Phoenix, Vitek 2, MicroScan) chosen as target index tests prior to any literature search activities; *Comparator* was the reference test (e.g., broth microdilution) used in the study, to which all included index tests were compared; *Outcome* included agreement measurements (categorical and essential) and error rates (very major errors and major errors) for the index tests, compared to the reference method.

### Search and selection criteria

Studies that involved AST, including retrospective, randomized and non-randomized prospective, as well as blinded and non-blinded designs, were retrieved. The index tests included in the search string for this study were Vitek 2, Phoenix, and MicroScan AST systems. The primary outcomes included: (1) categorical agreement (CA), calculated as: (number of isolates with the same susceptible, intermediate, or resistant results by the index test compared to reference) / (total number of isolates tested) X 100; (2) Essential agreement (EA): (number of minimum inhibitory concentration [MIC] results for isolate testing that are within +/- 1 doubling dilution as compared to reference result) / (total number of isolates tested) X 100; (3) very major errors (VMEs): (number of false susceptible results [by index test] / total number of resistant results [by reference method]) X 100; and (4) major errors (MEs): (number of false resistant [by index test] / total number of susceptible results [by reference method]) X 100.

Both MEDLINE (1946 to October 17, 2024) and Embase (1974 to October 17, 2024) electronic databases were searched with the following terms: (broth dilution/ or antibiotic sensitivity/ or minimum inhibitory concentration/ or antibiotic resistance/ or drug sensitivity.mp.) OR (microbial sensitivity tests/ or drug resistance, bacterial/ or anti-bacterial agents/ or drug resistance, microbial/) combined with the keywords “Vitek,” “Phoenix,” and “MicroScan” (**Table S1**). All retrieved articles were assessed for relevance in the first round of screening (Round 1) using predetermined inclusion/exclusion criteria. The inclusion criteria were: (1) articles with organisms that were identified and subsequently underwent AST; (2) articles with at least one of the following: CA, EA, VME rate, or ME rate as a performance outcome; (3) articles comparing at least two of the three target index tests with a common reference (e.g., broth microdilution); (4) articles involving a human study; (5) articles published in the English language. Secondary inclusion criteria were index performance results stratified by drug class, organism at the species or genus level, AST guideline (e.g., CLSI), and guideline year. The exclusion criteria were: (1) articles that were not peer-reviewed; (2) articles with an unclear or indistinct research question; (3) articles without performance data specific to AST; (4) articles that did not identify index or reference tests used or did not include information about the drugs utilized or organisms tested; (5) articles without a common reference to which two or more of the target index tests were compared; (6) articles in which data were collected in an unethical manner; (7) articles whose data could not be extracted in a way that supported analytical requirements. Secondary exclusion criteria were articles not in the English language and articles that did not involve humans.

Full text reviews were conducted on articles that passed the initial screening (Round 1) to identify those meeting the inclusion/exclusion criteria based on study methodology, data type (e.g., agreement, error rates), and data format (e.g., raw data, only point estimates and 95% confidence intervals [95% CI]). In this second round of assessment (Round 2), data extraction tables were prepared to document study characteristics and to record raw data. A modified Newcastle-Ottawa Scale was used to evaluate the risk of bias (individual study quality),^16^ which included the following bias domains: detection (measurement of test result), reporting (failure to adequately control confounding, failure to measure all known prognostic factors), and spectrum (eligibility criteria, formation of cohort, selection of participants). Risk of bias summary assessments for individual studies were categorized as high, moderate, or low. The overall quality of evidence for the risk estimate outcomes (all included studies) was obtained using a modified Grading of Recommendations, Assessment, Development and Evaluation (GRADE) methodology for observational diagnostic studies.^17^

The seven domains used to ascertain the overall study quality and strength across the four outcomes (CA, EA, VME, and ME) were: 1) Confounder effect, 2) Consistency, 3) Directness, 4) Magnitude of effect, 5) Precision, 6) Publication bias, and 7) Risk of bias. For all seven domains, the articles were assigned a score, as follows: high quality (score = 0), moderate quality (score = 0.5), or low quality (score = 1.0). The scores of the articles, matched to their inclusion in any of the four outcomes, were then averaged and the articles were categorized as follows: high quality (green) when 0≤X≤0.33; moderate quality (yellow) when 0.33<X≤0.66; low quality (red) when 0.66<X≤1.00). An unclear categorization was used when insufficient information was available to confidently assign a quality rating for one or more domains, such as missing methodological details or lack of transparency in reporting. For risk of bias, three subdomains were utilized: detection, performance, and spectrum bias. The overall bias was calculated as the average of the three subdomain scores. Overall quality, based on the distribution of green, yellow, red, and unclear classifications, was assigned according to the overall quality rating schedule shown in **Table S2**.

### Data analysis

Data extraction was performed by two reviewers and discrepancies were adjudicated by a third reviewer. An author who was not involved in screening and data extraction performed all statistical analysis using R software (version 4.5.2)^18^ along with the meta^19^ and metaphor^20^ packages. A random effects model was used to estimate the rate ratio and 95% confidence interval for each comparison. The same method was applied to subgroup meta-analyses; subgroups were defined by Gram stain, antibiotic utilized, guideline and guideline year, VME % threshold (≥ 3%), and reference method. Funnel plots were generated to look for publication bias or study quality issues and forest plots were used to display results. The minimum number of studies required for synthesis was n=3.

## RESULTS

Overall, 431 records (91% of total articles retrieved before deduplication) were obtained from literature searches conducted using MEDLINE and Embase databases (in Ovid), while 44 records (9% of total) were obtained through handsearching of literature cited in relevant retrieved articles and through searches of RightFind® or internal databases. Deduplication in Ovid (MEDLINE and Embase) removed 151 records, and 5 additional duplicates not identified by Ovid were also removed manually prior to title and abstract screening. Thus, 319 records (275 from MEDLINE/Embase and 44 retrieved through handsearching) were carried into Round 1 of the literature review (title and abstract screening). Of the total 319 articles assessed, 171 were excluded (reasons are listed in Figure 1). The remaining 148 records were carried into Round 2 (full text screening), at which stage 109 additional records were excluded. Data extraction and meta-analysis were performed on 39 articles— 34 (87%) from MEDLINE/Embase and 5 (13%) from handsearch (**Figure 1**).

**FIGURE 1.**
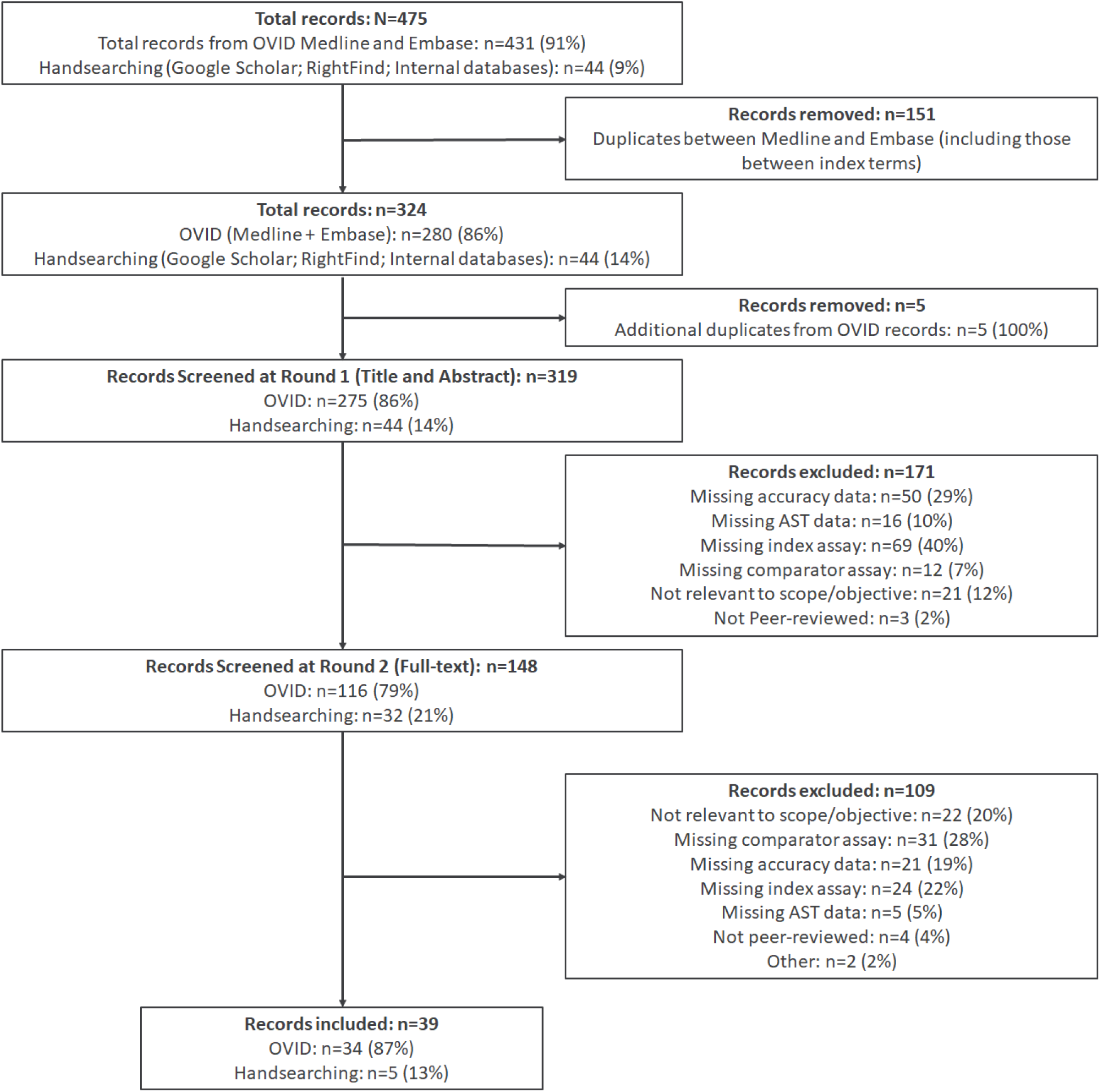
Reconciliation though two screening rounds of source articles retrieved from the systematic literature review of MEDLINE and Embase databases through the Ovid portal. **Abbreviation:** AST, antimicrobial susceptibility testing

The data set derived from the 39 included articles encompassed more than 20 distinct genera, over 35 species categories, more than 25 drug classes, and over 40 individual drugs, all related to AST. Of the 39 articles, 14 (35.9%) included prospectively collected clinical specimens while 25 (64.1%) included stored specimens — either clinical specimens (n=15; 38.5%) or characterized isolates (n=10; 25.6%) **(Table 1).** Reference testing methods differed across studies; the reference types included broth microdilution/previously characterized reference isolates (reference strain) (n=31 articles; 79.5%), agar dilution (n=5 articles; 12.8%), consensus comparator (n=2 articles; 5.1%), and mecA/mecC PCR (n=1 article; 2.6%). One article utilized a consensus approach that included three methods (broth microdilution, agar dilution, and disc diffusion).^21^ One study employed a rotating composite reference method in which each of the three individual index tests was compared against a consensus result (agreement between 2 out of 3 index tests).^22^ Twelve (12) of the 31 articles (38.7%) that reported broth microdilution or reference strain as the reference method indicated that AST reagents were stored frozen, one article (3.2%) reported lyophilization of reference reagents, and 18 (58.1%) did not indicate reagent storage conditions. In total, 22 (56.4%) articles represented studies conducted across multiple testing sites while the remaining 17 articles (43.6%) were from single-site studies. By country, 17 (43.6%) studies were conducted in the United States while the remainder (22; 56.4%) were either conducted outside the United States or included the United States along with at least one other country (**Table 1**).

**Table 1.**
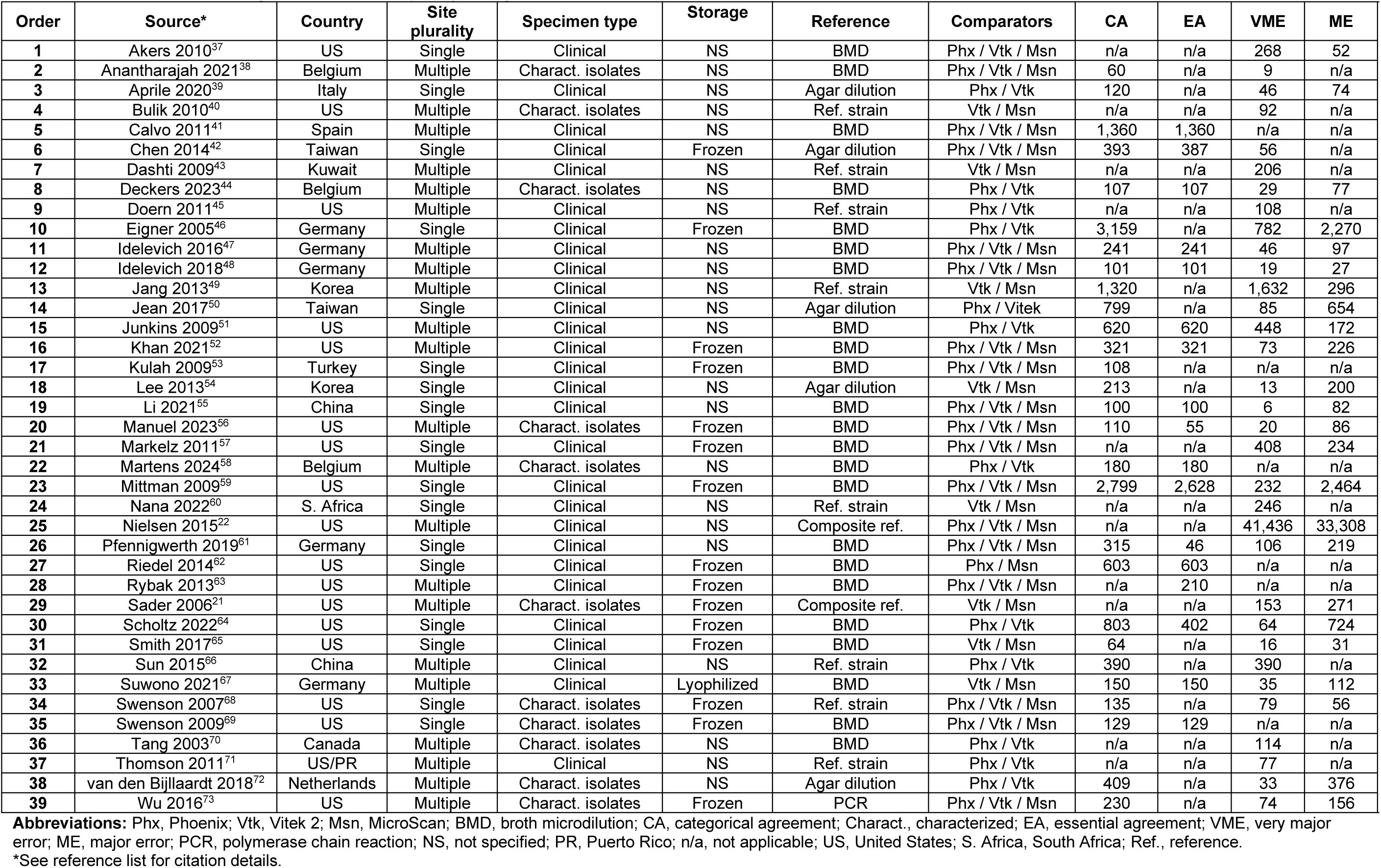
Information characterizing antimicrobial susceptibility testing for each article mentioned in this report.

Among the 39 included articles, comparisons involving two or more AST systems (Phoenix, Vitek 2, and/or MicroScan) to a common reference were reported in 28 articles for CA, 17 for EA, 33 for VME, and 24 for ME. Among studies evaluating groups of more than 1,000 specimens, 14.3% (4/28) reported CA, 11.8% (2/17) reported EA, 6.1% (2/33) reported VME, and 12.5% (3/24) reported ME. For studies with groups of 100–1,000 specimens, 78.6% (22/28) reported CA, 76.5% (13/17) reported EA, 36.4% (12/33) reported VME, and 50.0% (12/24) reported ME. In studies with groups of fewer than 100 specimens, 7.1% (2/28) reported CA, 11.8% (2/17) reported EA, 57.6% (19/33) reported VME, and 37.5% (9/24) reported ME (**Table 1**).

Most studies were associated with low or moderate detection bias (79.5%; 31/39), performance bias (100%; 39/39), and spectrum bias (92.3%; 36/39) (**Figure S1**). Quality of evidence was likely affected by confounder effect caused by factors such as incubation conditions, technical variability, inoculum size, and growth media, which may differ across laboratories separated by region or country. In addition, publication bias likely influenced the quality of evidence for the meta-analyses to some extent, although funnel plot analyses suggest that the observed standard error may be largely attributable to study heterogeneity (**Figure 2**). Overall, the quality of evidence was rated as moderate across all four outcomes of interest (CA, EA, VME, and ME) (**Table 2**).

**FIGURE 2.**
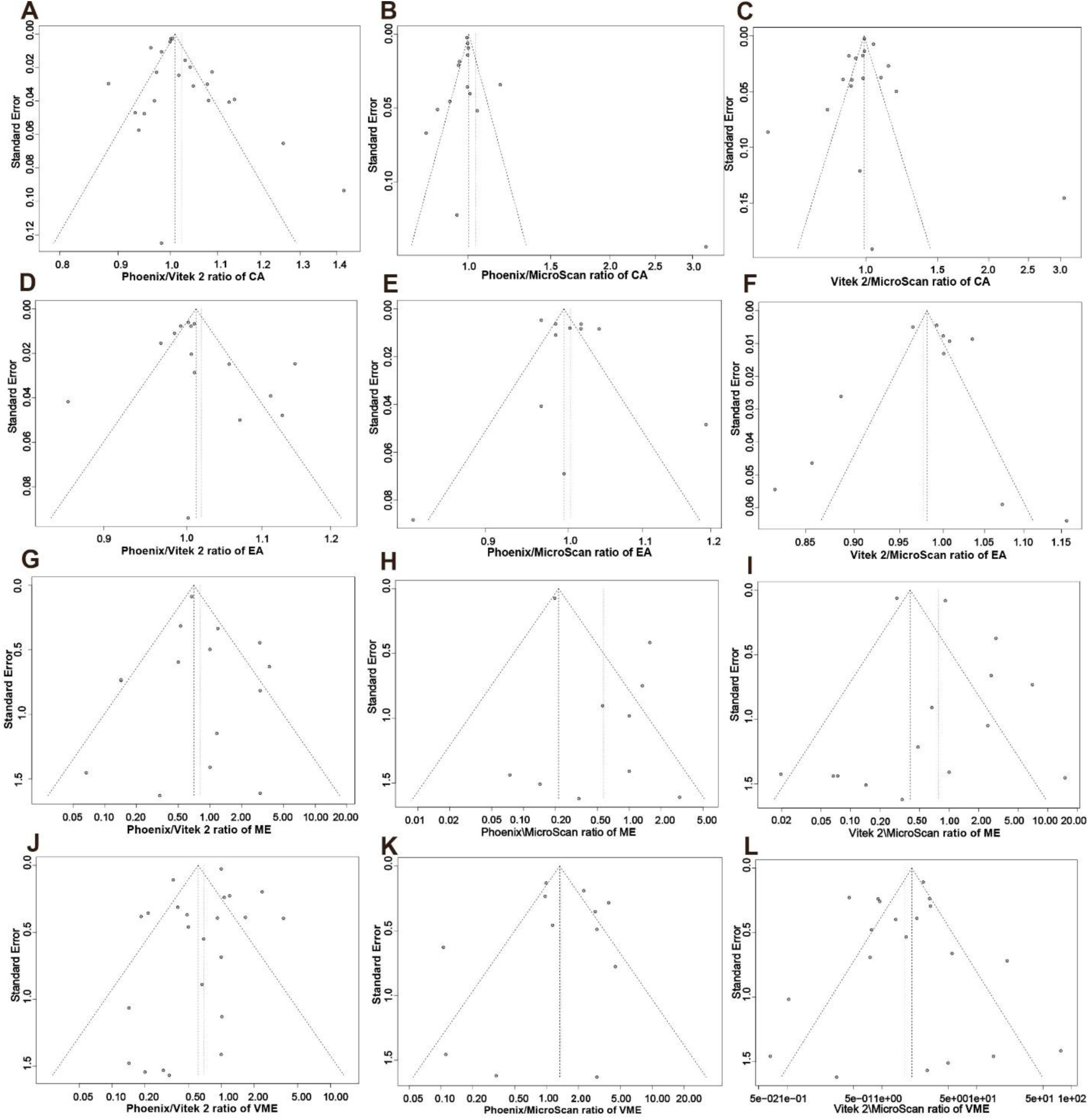
Funnel plots of index test ratios [x-axis] versus standard error [y-axis]. Funnel plots are indicative of publication bias, with those plotted circles falling outside the “funnel” as potentially reflecting more bias compared to those inside the funnel (inside largely reflecting heterogeneity of studies). **Abbreviations:** EA, essential agreement; CA, categorical agreement; VME, very major error; ME, major error.

**Table 2.** Overall quality of evidence for outcomes (modified GRADE)^17^.

| Outcome | No. Studies | Total N | Confounder | Consistency | Directness | Magnitude of effect | Precision | Publication bias | Risk of bias | Overall |
| --- | --- | --- | --- | --- | --- | --- | --- | --- | --- | --- |
| CA | 28 | 15,339 | Moderate | Moderate | High | Low | Low | Moderate | Low | Moderate |
| EA | 17 | 7,640 | Moderate | Moderate | High | Low | Low | Moderate | Low | Moderate |
| VME | 33 | 47,401 | Moderate | Moderate | High | Moderate | Moderate | Moderate | Low | Moderate |
| ME | 24 | 42,264 | Moderate | Moderate | High | Moderate | Moderate | Moderate | Low | Moderate |

Abbreviations: GRADE, Grading of Recommendations, Assessment, Development and Evaluation; CA, categorical agreement; EA, essential agreement; VME, very major error; ME, major error

Raw data were extracted and then rate ratios were calculated. For meta-analyses, results were reported as rate ratios of two index test results; thus, ratios were reported for the comparisons performed that involved Phoenix and/or Vitek 2 and/or MicroScan. The number of studies for overall results (no stratification of data for rate ratios related to CA, EA, VME, and ME) ranged from 11 to 25 and results of analyses are shown in **Table 3**. No statistically significant differences were obtained for either CA or EA when performing rate ratio analyses for the index tests (**Table 3**; **Figure S2**; **Figure S3**). Overall, across all organism morphologies and breakpoint applications, Phoenix had fewer VMEs than Vitek 2 (rate ratio of Phoenix-to-Vitek 2 = 0.69 [95% CI: 0.47, 1.02]), although the difference was only marginally significant (*p*=0.062). In contrast, Vitek 2 had significantly more VMEs than MicroScan (rate ratio of Vitek 2-to-MicroScan = 1.74 [95% CI: 1.01, 2.99]; *p*=0.045); the rate ratio of Phoenix-to-MicroScan was 1.36 [95% CI: 0.73, 2.56] (*p*=0.332) **(Table 3 and Figure 3)**. The rate ratio point estimates for overall ME were 0.81 for Phoenix versus Vitek 2, 0.57 for Phoenix versus MicroScan, and 0.78 for Vitek 2 versus MicroScan (**Table 3; Figure S4**). In all three comparisons, the 95% CIs spanned 1.0 and none of the differences were statistically significant.

**FIGURE 3.**
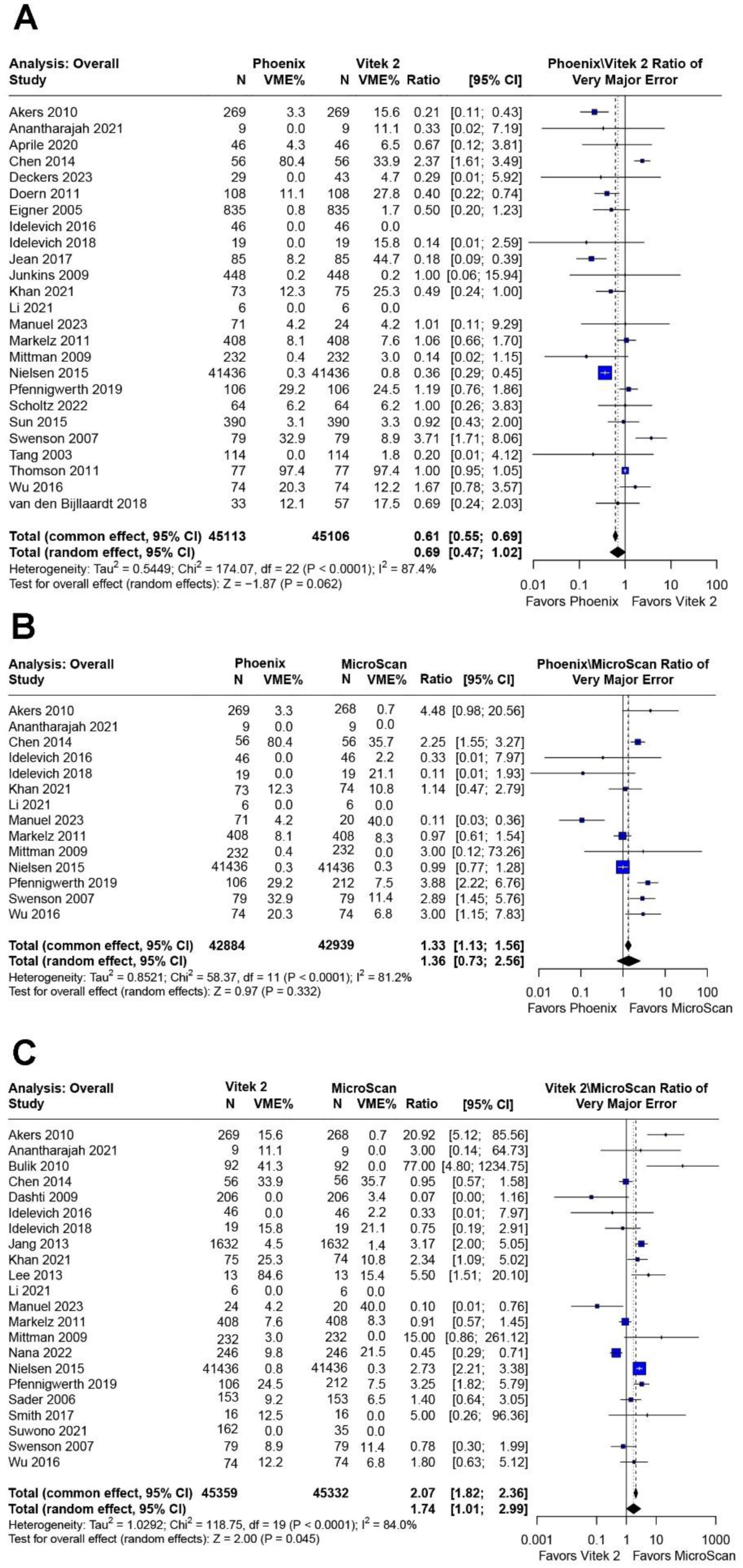
Forest plots for Index X / Index Y (X and Y represent Phoenix and/or Vitek 2 and/or MicroScan) ratios following conversion to weighted averages and statistical calculation for common and random effects. Overall analyses, which included any article reporting VME values for ≥2 instruments (with a common reference method), are shown. Forest plots for Index X / Index Y (X and Y represent Phoenix and/or Vitek 2 and/or MicroScan) very major error ratios following conversion to weighted averages and statistical calculation for common and random effects. The bars spanning from each of the square point estimates represent confidence intervals. The size of the square plots represents the relative size of the group number for each included study. The threshold for statistical significance was *p*<0.05. (A) (B) and (C) represent comparisons for Phoenix versus Vitek 2, Phoenix versus MicroScan, and Vitek 2 versus MicroScan, respectively. **Abbreviation:** VME, very major error

**Table 3.** Antimicrobial susceptibility testing outcomes from included articles for Phoenix, Vitek 2, and MicroScan—overall analysis.

| Index 1 | Index 2 | No. Studies | Outcome | Rate 1<sup>a</sup> | Rate 2<sup>a</sup> | Ratio<sup>b</sup> | p-value |
| --- | --- | --- | --- | --- | --- | --- | --- |
| Phoenix | Vitek 2 | 23 | CA | 12,402/13,244 (93.64%) | 12,656/13,607 (93.01%) | 1.02 [0.99, 1.06] | 0.148 |
| Phoenix | MicroScan | 15 | CA | 6,746/7,259 (92.93%) | 7,897/8,474 (93.19%) | 1.04 [0.91, 1.19] | 0.571 |
| Vitek 2 | MicroScan | 18 | CA | 7,748/8,608 (90.01%) | 8,214/9,015 (91.11%) | 1.00 [0.88, 1.14] | 0.949 |
| Phoenix | Vitek 2 | 15 | EA | 6,880/7,307 (94.16%) | 6,516/7,011 (92.94%) | 1.02 [0.99, 1.05] | 0.260 |
| Phoenix | MicroScan | 11 | EA | 6,047/6,367 (94.97%) | 7,326/7,651 (95.75%) | 1.00 [0.98, 1.03] | 0.766 |
| Vitek 2 | MicroScan | 11 | EA | 5,776/6,165 (93.69%) | 6,300/6,595 (95.53%) | 0.98 [0.93, 1.02] | 0.333 |
| Phoenix | Vitek 2 | 25 | VME | 410/45,113 (0.91%) | 671/45,106 (1.49%) | 0.69 [0.47, 1.02] | 0.062 |
| Phoenix | MicroScan | 14 | VME | 286/42,884 (0.67%) | 222/42,939 (0.52%) | 1.36 [0.73, 2.56] | 0.332 |
| Vitek 2 | MicroScan | 22 | VME | 641/45,359 (1.41%) | 317/45,332 (0.70%) | 1.74 [1.01, 2.99] | 0.045 |
| Phoenix | Vitek 2 | 19 | ME | 299/42,635 (0.70%) | 435/41,902 (1.04%) | 0.81 [0.46, 1.40] | 0.443 |
| Phoenix | MicroScan | 12 | ME | 242/38,287 (0.63%) | 1,134/38,364 (2.96%) | 0.57 [0.26, 1.23] | 0.154 |
| Vitek 2 | MicroScan | 17 | ME | 533/38,219 (1.39%) | 1,323/39,274 (3.37%) | 0.78 [0.34, 1.77] | 0.551 |

**Abbreviations:** CA, categorical agreement; EA, essential agreement; ME, major error; VME, very major error
<sup>a</sup>Rate reported here is based on raw data
<sup>b</sup>Ratio is based on random effects modeling

As VME was the only outcome for which statistically significant differences were demonstrated in the overall meta-analysis, further stratification and subgroup analyses were performed with VME as the outcome of interest. Data were stratified by Gram stain (gram-positive versus gram-negative) and by guideline (CLSI versus EUCAST). The number of studies following stratification by either Gram stain or guideline ranged from 4 to 18 and results are shown in **Table 4**. For gram-negative organisms, Phoenix had significantly fewer VMEs than Vitek 2 (rate ratio = 0.53 [0.36, 0.77]; *p*<0.001), while Phoenix and MicroScan had similar VME rates (rate ratio = 0.96 [0.38, 2.41]; *p*=0.929). For gram-negative organisms, the rate ratio for Vitek 2-to-MicroScan was 1.91 [0.94, 3.92], although the difference was only approaching significance (*p*=0.076). No significant differences in VME rates were associated with gram-positive organisms for any of the instrument comparisons, although Phoenix had a marginally significant higher rate than MicroScan (rate ratio = 1.84 [0.92, 3.68]; *p*=0.086). Stratification by articles that reported VME rates according to CLSI guidelines revealed no significant differences for any of the instrument comparisons. However, Vitek 2 had a marginally significant higher VME rate than MicroScan (rate ratio = 1.80 [0.97, 3.36]; *p*=0.064). No differences in VME rates were observed for data stratified by EUCAST guidelines for any of the instrument comparisons (**Table 4**).

**Table 4.** Antimicrobial susceptibility testing outcomes from included articles for Phoenix, Vitek 2, and MicroScan—VME across analyses for subgroups.

| Index 1 | Index 2 | No. Studies | Subgroup Analysis | Rate 1<sup>a</sup> | Rate 2<sup>a</sup> | Ratio<sup>b</sup> | p-value |
| --- | --- | --- | --- | --- | --- | --- | --- |
| Phoenix | Vitek 2 | 18 | gram-negative | 274/25,522 (1.08%) | 576/25,515 (2.26%) | 0.53 [0.36, 0.77] | <0.001 |
| Phoenix | Vitek 2 | 9 | gram-positive | 134/19,591 (0.68%) | 91/19,591 (0.46%) | 1.41 [0.82, 2.40] | 0.211 |
| Phoenix | Vitek 2 | 18 | CLSI | 341/44,570 (0.77%) | 596/44,540 (1.34%) | 0.68 [0.43, 1.09] | 0.107 |
| Phoenix | Vitek 2 | 7 | EUCAST | 64/378 (16.93%) | 70/416 (17.41%) | 1.01 [0.75, 1.36] | 0.962 |
| Phoenix | MicroScan | 10 | gram-negative | 158/24,235 (0.65%) | 127/24,290 (0.52%) | 0.96 [0.38, 2.41] | 0.929 |
| Phoenix | MicroScan | 5 | gram-positive | 128/18,649 (0.69%) | 95/18,649 (0.51%) | 1.84 [0.92, 3.68] | 0.086 |
| Phoenix | MicroScan | 9 | CLSI | 223/42,403 (0.53%) | 165/42,366 (0.39%) | 1.53 [0.85, 2.77] | 0.159 |
| Phoenix | MicroScan | 5 | EUCAST | 60/316 (18.99%) | 51/422 (12.09%) | 1.01 [0.25, 4.10] | 0.994 |
| Vitek 2 | MicroScan | 18 | gram-negative | 558/26,710 (2.09%) | 222/26,683 (0.83%) | 1.91 [0.94, 3.92] | 0.076 |
| Vitek 2 | MicroScan | 5 | gram-positive | 83/18,649 (0.45%) | 95/18,649 (0.51%) | 0.89 [0.62, 1.27] | 0.519 |
| Vitek 2 | MicroScan | 16 | CLSI | 521/43,793 (1.19%) | 248/43,786 (0.57%) | 1.80 [0.97, 3.36] | 0.064 |
| Vitek 2 | MicroScan | 7 | EUCAST | 92/1,370 (6.72%) | 63/1,349 (4.67%) | 1.66 [0.87, 3.19] | 0.125 |

**Abbreviations:** VME, very major error; CLSI, Clinical and Laboratory Standards Institute; EUCAST, European Committee on Antimicrobial Susceptibility Testing
<sup>a</sup>Rate reported here is based on raw data
<sup>b</sup>Ratio is based on random effects modeling

VME results were stratified to determine how specific drug classes contributed to overall Vitek 2 VMEs. MicroScan had fewer VMEs associated with aminoglycosides, monobactams, cephems, macrolides, and tetracyclines, although the differences were only significant for aminoglycosides and monobactams (**Figure S5**). Removal of aminoglycosides (the drug class associated with the highest Vitek 2 VME rate) from overall VME analysis rendered the VME ratio for MicroScan compared to Vitek 2 close to the value of one (rate ratio = 1.46 [0.93; 2.29]; *p*=0.099) (**Figure S6**).

While stratification by antimicrobial class provided insight into drug-specific contributions to VME rates, test performance may also be influenced by other factors, including the breakpoints applied during susceptibility interpretation. Automated AST systems must be regularly updated to reflect current interpretive criteria established by organizations such as CLSI, EUCAST, or FDA. In the United States, College of American Pathologists (CAP)-accredited laboratories were required to adopt breakpoints with at least the 2021 standards by January 1, 2024.^23^ Given the clinical importance of accurate susceptibility reporting and the regulatory requirement for laboratories to align with current interpretive standards, we also examined whether differences in breakpoint application could account for variability in VME rate across systems. This approach allowed us to assess how system performance trends have evolved over time in relation to shifting interpretive standards.

In **Figure 4**, VME ratios for Phoenix vs MicroScan, Phoenix vs Vitek 2, and Vitek 2 vs MicroScan are plotted as a function of CLSI guideline year. Phoenix-to-Vitek 2 VME ratios remained relatively flat (and below the value of 1.0) across all time points. The Vitek 2-to-MicroScan VME ratio decreased steadily from 2010 to 2020, whereas the Phoenix-to-MicroScan ratio, except for a small increase between 2010 and 2015, also decreased steadily until 2020.

**FIGURE 4.**
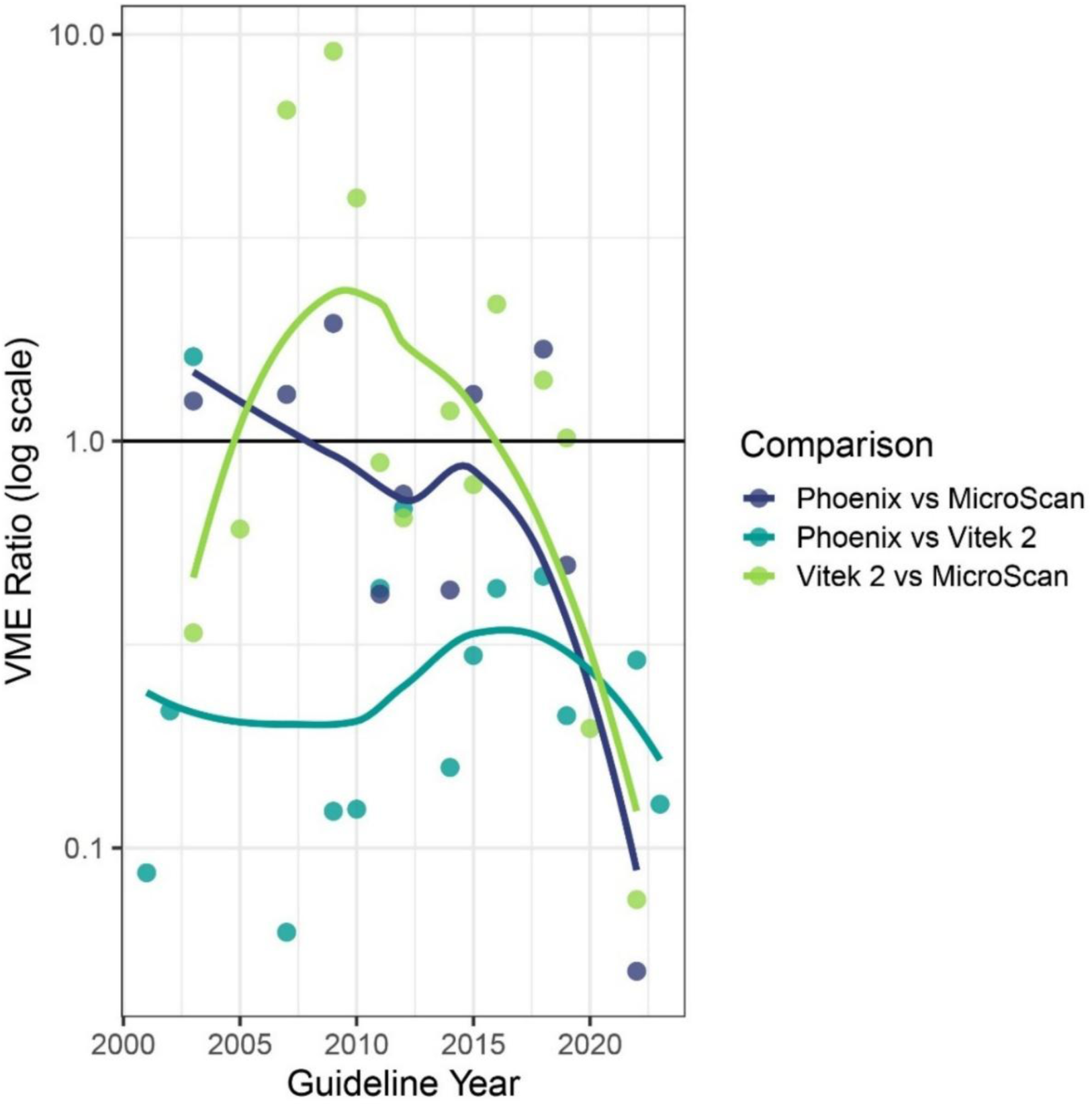
Plots of VME ratio (log scale; y-axis) over different years of CLSI guidelines (x-axis). Individual plots (colored circles) represent articles reporting Vitek 2 - MicroScan values (green), Phoenix - MicroScan values (dark blue), or Phoenix - Vitek 2 values (light blue). The first instrument listed in the key acts as the numerator of the comparative ratio values (for example, Vitek 2 versus MicroScan puts Vitek 2 in the numerator). **Abbreviation:** VME, very major error

Three percent (3%) is a consensus threshold VME rate that is acceptable for detection of resistant strains during AST, as outlined by CLSI guidelines.^24,25^ Stratifying the data set according to <3% (acceptable) and ≥3% (unacceptable) allowed for analysis of clinically and operationally relevant benchmarks. This threshold is particularly important for laboratory directors and quality managers, as it helps assess whether AST platforms are performing within acceptable limits for patient safety and regulatory expectations.^26^ The overall VME rates for the three platforms were stratified using two approaches. The first approach analyzed the data by antibiotic/organism combination for each article. When doing so, MicroScan, Phoenix, and Vitek 2 were associated with 129, 155, and 195 antibiotic/organism combinations, respectively (**Figure 5**). MicroScan had 77.5% (100/129) antibiotic/organism combinations with acceptable VME rates whereas Phoenix and Vitek 2 had 76.1% (118/155) and 65.1% (127/195), respectively (**Figure 5**). A significant p-value, 0.02, for the Chi-square test indicates that platform and acceptable VME rates are associated. However, no significant differences were found for post hoc pair-wise comparisons using the Bonferroni adjusted alpha. This analysis was complemented by a drug class-level view of VME acceptability across published studies, illustrating the proportion of articles in which all reported values remained below the 3% threshold for each platform (**Figure S7**).

**FIGURE 5.**
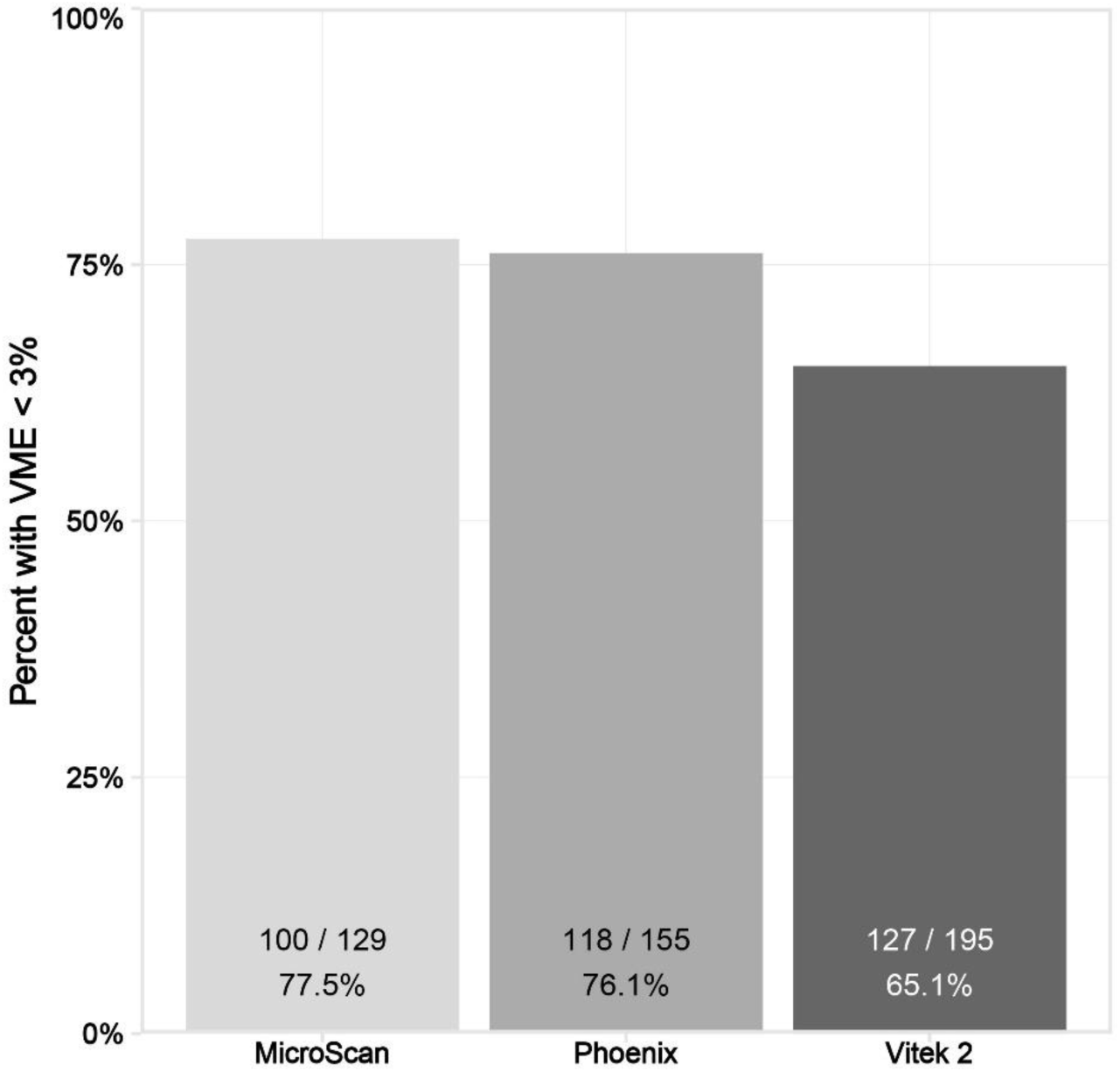
Antibiotic/organism combinations within articles/studies (regardless of antibiotic/organism combination) reporting VME were assessed for any VME value ≥3%. Here antibiotic/organism combinations with no VME values ≥3% are plotted as a percentage of total along the x-axis across MicroScan, Phoenix, and Vitek 2 platforms. **Abbreviations:** VME, very major error; ns, not significant

A subset of five studies employed agar dilution, which may also have introduced variability due to differences in technique and interpretation (see Table 1). To explore how these methodological differences may have influenced overall VME estimates, we performed sensitivity analyses excluding studies that used agar dilution (**Figure S8**) as well as the Nielsen study (**Figure S9**). These analyses clarify how reference method variability may have influenced the observed differences in VME rates across platforms. The resulting VME ratios were directionally consistent with the overall analysis, with no substantial shifts in comparative trends across platforms.

## DISCUSSION

To our knowledge, this is the first systematic literature review and meta-analysis to report CA, EA, VME, and ME values of multiple commercial AST systems to compare their relative performance. Overall, the three commercial AST systems performed well and did not show significant differences in CA (all ≥90%) and EA (all >92%). ME rates were low overall, although MicroScan showed higher ME percentages in the comparisons of Phoenix-to-MicroScan (2.96%) and Vitek 2-to-MicroScan (3.37%). Vitek 2 showed higher overall VME rates than MicroScan (*p*=0.045) and Phoenix (marginally significant difference; *p*=0.062). This finding is particularly important as VMEs—false susceptibility results—can lead to inappropriate treatment decisions, including the administration of unwarranted antibiotics. Such errors are particularly concerning in cases of high-risk infections, such as sepsis or bacteremia, where timely and accurate therapy is critical. Bartoletti et al. (2022) demonstrated that unwarranted therapy resulting from AST errors generated by semi-automated systems (Vitek 2 or MicroScan) for carbapenem-resistant *Enterobacterales* was associated with a 30% increase in 30-day mortality compared to effective (clinically warranted) therapy.^27^ Similarly, additional work has shown that delays or inaccuracies in susceptibility reporting can prolong infections, increase ICU stays, and drive up healthcare costs.^28^ From a public health perspective, persistent use of antibiotics, misaligned with the infectious etiology, not only fails to resolve infections but also promotes the selection and spread of multidrug-resistant organisms, further contributing to the global burden of AMR.^29^ As reported in the Lancet AMR series, estimates show that AMR is now associated with nearly 5 million deaths annually, with inappropriate antibiotic use often stemming from diagnostic uncertainty, which represents a key driver.^30^ These findings highlight the critical need for accurate and timely AST to guide effective therapy, reduce patient harm, and preserve the utility of existing antimicrobials.

Given the clinical and public health impact of VMEs, it is important not only to understand the difference in error rates across platforms but also to investigate underlying factors contributing to those differences. Subgroup analyses provided further insight into the nature of these performance differences. In comparison to MicroScan, Vitek 2 showed the highest VME rates in association with specific drug classes, including aminoglycosides, monobactams, cephems, macrolides, and tetracyclines (**Figure S5**). These drug classes comprise a significant portion of the antimicrobial agents used in clinical practice, particularly for the treatment of gram-negative infections. Cephalosporins and macrolides are frequently employed as first-line or empiric therapies for common infections such as community-acquired pneumonia, urinary tract infections, and skin and soft tissue infections.^31,32^ Aminoglycosides and monobactams, while often reserved for more severe or drug-resistant infections, are critical components of combination regimens for hospital-acquired infections, sepsis, and infections caused by multidrug-resistant organisms.^33,34^ Inaccurate susceptibility results for these agents can lead to inappropriate or delayed therapy, increased risk of treatment failure, and adverse clinical outcomes, particularly in critically ill or immunocompromised patients.^35^ The inclusion of these drug classes appears to have substantially contributed to the observed differences in VME rates within this meta-analysis. When these classes were excluded from the analysis, VME rates across all three systems appeared more closely aligned (**Figure S6**), suggesting that drug-specific factors may meaningfully influence overall system performance. While Vitek 2 exhibited reduced VME rates for selected drug classes compared to Phoenix and MicroScan, these differences did not have a major impact on overall VME rate due to the sample sizes associated with those classes (**Figure S7**; **Table 3**). In other subgroup analyses, both Phoenix and MicroScan showed lower VME rates for AST associated with gram-negative organisms. However, Vitek 2 showed either no difference (versus MicroScan),or lower VME rates (versus Phoenix) for AST in association with gram-positive organisms, although not statistically significant (**Table 4**).

Another potential factor that could influence differential instrument performance is the use of extrapolated results. In the Vitek 2 system, MIC values are not directly measured across the full dilution range; instead, susceptibility is inferred through algorithmic extrapolation when certain concentrations are skipped on the testing panel. While this approach can streamline testing and reduce AST panel or card complexity, it may introduce error—particularly when the skipped concentrations are near clinical breakpoints or when resistance mechanisms produce non-linear MIC shifts. The extent to which extrapolated results contributed to the observed VME rates is unclear, as no studies in this analysis distinguished between directly measured and inferred values. Nonetheless, these findings highlight the importance of considering both drug-specific performance and interpretive methodologies—such as extrapolation—when evaluating AST system accuracy.

The evolving nature of breakpoint guidelines adds another layer of complexity to AST system performance. As interpretive criteria are updated by organizations such as CLSI and EUCAST, AST systems must adapt to maintain clinical relevance. These updates can significantly alter susceptibility categorizations, and systems that do not incorporate them promptly may yield outdated or misleading results. Simner et al. (2022) emphasized the importance of using current breakpoints to avoid misclassification of resistance.^36^ Although this meta-analysis did not explicitly assess the speed at which manufacturers implement updates, shifts in VME rates observed across studies conducted in different timeframes may reflect, at least in part, how breakpoint changes influence system performance. These findings suggest that AST accuracy is not a fixed attribute, but one that may fluctuate as interpretive standards evolve.

These considerations also underscore the important role of clinical laboratories in maintaining AST accuracy as breakpoints evolve. While manufacturers are required to validate AST system performance with each breakpoint update, clinical laboratories are only required to verify that MIC results are interpreted correctly according to the new breakpoints. This distinction is critical because interpretive accuracy does not necessarily ensure that MIC values remain reliable as breakpoints shift. Our findings suggest that performance characteristics—particularly VME rates—can shift over time, highlighting the potential value of more comprehensive verification practices at the laboratory level. As breakpoint updates become more frequent and nuanced, laboratories may need to consider more robust validation strategies, such as periodic MIC correlation studies with reference methods or additional verification for results near clinical breakpoints, to ensure continued accuracy and patient safety.

Notably, in certain contexts, all three systems exceeded the 3% VME threshold—a benchmark frequently referenced in regulatory and clinical guidance.^24,25^ This observation highlights that even widely used FDA-cleared systems may occasionally fall short of optimal performance, particularly when challenged by evolving breakpoints or complex antibiotic/organism combinations. These findings reinforce the importance of ongoing post-market surveillance and real-world performance monitoring. Ongoing analyses at the genus/species level and by antimicrobial class may provide more granular insights into system-specific strengths and weaknesses, which could inform targeted quality assurance efforts.

## LIMITATIONS

There are important limitations to consider when interpreting the findings of this study. Although broth microdilution is widely regarded as the gold standard for antimicrobial susceptibility testing, not all studies used this method exclusively. Differences in reference methods across included studies may have impacted the accuracy of endpoints included in this meta-analysis, particularly where comparator methods varied. For example, the article by Nielsen et al (2015), which contributed to a relatively large percentage of the overall data for VMEs, employed a reference method involving a 2-out-of-3 consensus result for each index test as opposed to a single reference method (e.g., broth microdilution).^22^ In addition, a small number of studies employed agar dilution as the reference method, which may introduce variability in performance comparisons. Additionally, among those studies that did use broth microdilution, it was often unclear whether frozen or lyophilized panels were used.

Breakpoint utilization also differed by year and guideline (e.g., CLSI versus EUCAST), which directly affected susceptibility categorization and contributed to inconsistencies in reported performance. Although one of the most striking and informative aspects of this meta-analysis was observing how system performance varied over time as breakpoints evolved, this same variability must also be acknowledged as a limitation. Because studies applied different breakpoints depending on when they were conducted, the analysis does not allow for a direct, time-fixed comparison of system performance.

A substantial number of studies were excluded due to incomplete reporting—such as missing accuracy data, index methods, or comparators—which may limit the generalizability of the results. While subgroup analyses by Gram classification and drug class were conducted, species-level and antimicrobial-specific analyses were constrained by limited sample sizes, reducing the ability to draw more detailed conclusions about system-specific performance.

## CONCLUSIONS

This meta-analysis demonstrated that Phoenix, Vitek 2, and MicroScan AST systems generally showed similar overall agreement with reference methods. However, differences in very major error rates, particularly when stratified by organism type and drug class, highlight important nuances in system performance that may influence clinical reliability. These findings emphasize that AST system accuracy is not static and may be influenced by factors such as interpretive methodologies and evolving breakpoint criteria. As susceptibility criteria continue to change, both manufacturers and clinical laboratories must remain proactive in updating and verifying system performance. Ensuring the reliability of AST results in this dynamic landscape is critical to supporting timely, appropriate therapy and advancing antimicrobial stewardship efforts. Future studies should continue to evaluate the performance of updated AST panels and formulations, assess system responsiveness to evolving breakpoints, and investigate the real-world clinical impact of elevated VME rates—particularly in guiding appropriate antimicrobial therapy and informing stewardship strategies.

## Data Availability

Data produced in the present study are available upon reasonable request to the authors.

## ACKNOWLEDGEMENTS

We thank Hélène Tanguay, PhD, and Dorsey Mills, BA (both are employees of Waters Advanced Diagnostics, formerly BD Diagnostic Solutions) for their assistance with the investigation and literature screening phases of the study, as well as medical writing (HT) and editing support (DM). DM and HT have no other conflict of interest to disclose.

## AUTHOR CONTRIBUTION

KVB: Conceptualization, Validation, Formal analysis, Investigation, Data curation, Writing – Original draft, Manuscript – Review & Editing, Visualization, Supervision, Project administration.

LC: Conceptualization, Validation, Data curation, Manuscript – Review & Editing.

KK: Investigation, Manuscript – Review & Editing.

SK: Investigation, Manuscript – Review & Editing.

KB: Formal analysis, Data curation, Manuscript – Review and Editing, Visualization.

All authors approved the final manuscript and accept responsibility for its content.

## SUPLLEMENTAL INFORMATION

**Table S1.**
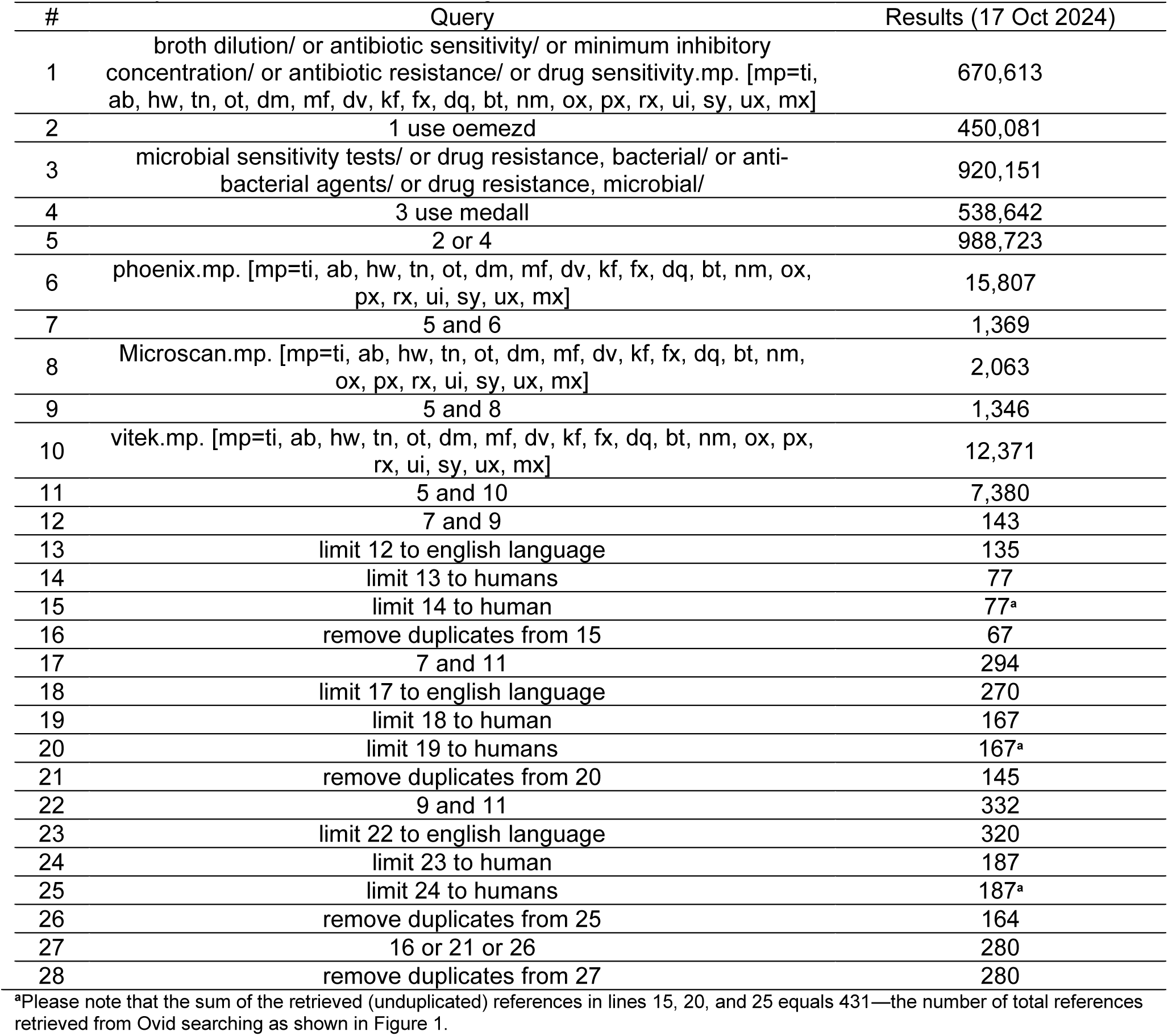
Systematic literature search string.

**Table S2.** Overall quality rating* schedule for included sources in this meta-analysis.

| #<br>Green | #<br>Yellow | #<br>Red | #<br>Unclear | Overall | #<br>Green | #<br>Yellow | #<br>Red | #<br>Unclear | Overall |
| --- | --- | --- | --- | --- | --- | --- | --- | --- | --- |
| 7 | 0 | 0 | 0 | High | 1 | 4 | 1 | 1 | Moderate |
| 6 | 1 | 0 | 0 | High | 1 | 4 | 0 | 2 | Moderate |
| 6 | 0 | 1 | 0 | High | 1 | 3 | 3 | 0 | Low |
| 6 | 0 | 0 | 1 | High | 1 | 3 | 2 | 1 | Low |
| 5 | 2 | 0 | 0 | High | 1 | 3 | 1 | 2 | Moderate |
| 5 | 1 | 1 | 0 | High | 1 | 3 | 0 | 3 | Moderate |
| 5 | 1 | 0 | 1 | High | 1 | 2 | 4 | 0 | Low |
| 5 | 0 | 2 | 0 | Moderate | 1 | 2 | 3 | 1 | Low |
| 5 | 0 | 1 | 1 | High | 1 | 2 | 2 | 2 | Moderate |
| 5 | 0 | 0 | 2 | High | 1 | 2 | 1 | 3 | Moderate |
| 4 | 3 | 0 | 0 | High | 1 | 2 | 0 | 4 | Low |
| 4 | 2 | 1 | 0 | Moderate | 1 | 1 | 5 | 0 | Low |
| 4 | 2 | 0 | 1 | High | 1 | 1 | 4 | 1 | Low |
| 4 | 1 | 1 | 1 | Moderate | 1 | 1 | 3 | 2 | Low |
| 4 | 1 | 2 | 0 | Moderate | 1 | 1 | 2 | 3 | Low |
| 4 | 1 | 0 | 2 | High | 1 | 1 | 1 | 4 | Low |
| 4 | 0 | 3 | 0 | Moderate | 1 | 1 | 0 | 5 | Low |
| 4 | 0 | 2 | 1 | Moderate | 1 | 0 | 6 | 0 | Low |
| 4 | 0 | 1 | 2 | High | 1 | 0 | 5 | 1 | Low |
| 4 | 0 | 0 | 3 | High | 1 | 0 | 4 | 2 | Low |
| 3 | 4 | 0 | 0 | Moderate | 1 | 0 | 3 | 3 | Low |
| 3 | 3 | 1 | 0 | Moderate | 1 | 0 | 2 | 4 | Low |
| 3 | 3 | 0 | 1 | Moderate | 1 | 0 | 1 | 5 | Low |
| 3 | 2 | 2 | 0 | Moderate | 1 | 0 | 0 | 6 | Low |
| 3 | 2 | 1 | 1 | Moderate | 0 | 7 | 0 | 0 | Moderate |
| 3 | 2 | 0 | 2 | Moderate | 0 | 6 | 1 | 0 | Moderate |
| 3 | 1 | 3 | 0 | Low | 0 | 6 | 0 | 1 | Moderate |
| 3 | 1 | 2 | 1 | Moderate | 0 | 5 | 2 | 0 | Moderate |
| 3 | 1 | 1 | 2 | Moderate | 0 | 5 | 1 | 1 | Moderate |
| 3 | 1 | 0 | 3 | Moderate | 0 | 5 | 0 | 2 | Moderate |
| 3 | 0 | 4 | 0 | Low | 0 | 4 | 3 | 0 | Moderate |
| 3 | 0 | 3 | 1 | Low | 0 | 4 | 2 | 1 | Moderate |
| 3 | 0 | 2 | 2 | Moderate | 0 | 4 | 1 | 2 | Moderate |
| 3 | 0 | 1 | 3 | Moderate | 0 | 4 | 0 | 3 | Moderate |
| 3 | 0 | 0 | 4 | Moderate | 0 | 3 | 4 | 0 | Low |
| 2 | 5 | 0 | 0 | Moderate | 0 | 3 | 3 | 1 | Low |
| 2 | 4 | 1 | 0 | Moderate | 0 | 3 | 2 | 2 | Moderate |
| 2 | 4 | 0 | 1 | Moderate | 0 | 3 | 1 | 3 | Moderate |
| 2 | 3 | 2 | 0 | Moderate | 0 | 3 | 0 | 4 | Low |
| 2 | 3 | 1 | 1 | Moderate | 0 | 2 | 5 | 0 | Low |
| 2 | 3 | 0 | 2 | Moderate | 0 | 2 | 4 | 1 | Low |
| 2 | 2 | 3 | 0 | Low | 0 | 2 | 3 | 2 | Low |
| 2 | 2 | 2 | 1 | Moderate | 0 | 2 | 2 | 3 | Low |
| 2 | 2 | 1 | 2 | Moderate | 0 | 2 | 1 | 4 | Low |
| 2 | 2 | 0 | 3 | Moderate | 0 | 2 | 0 | 5 | Low |
| 2 | 1 | 4 | 0 | Low | 0 | 1 | 6 | 0 | Low |
| 2 | 1 | 3 | 1 | Low | 0 | 1 | 5 | 1 | Low |
| 2 | 1 | 2 | 2 | Moderate | 0 | 1 | 4 | 2 | Low |
| 2 | 1 | 1 | 3 | Moderate | 0 | 1 | 3 | 3 | Low |
| 2 | 1 | 0 | 4 | Low | 0 | 1 | 2 | 4 | Low |
| 2 | 0 | 5 | 0 | Low | 0 | 1 | 1 | 5 | Low |
| 2 | 0 | 4 | 1 | Low | 0 | 1 | 0 | 6 | Low |
| 2 | 0 | 3 | 2 | Low | 0 | 0 | 7 | 0 | Low |
| 2 | 0 | 2 | 3 | Low | 0 | 0 | 6 | 1 | Low |
| 2 | 0 | 1 | 4 | Low | 0 | 0 | 5 | 2 | Low |
| 2 | 0 | 0 | 5 | Low | 0 | 0 | 4 | 3 | Low |
| 1 | 6 | 0 | 0 | Moderate | 0 | 0 | 3 | 4 | Low |
| 1 | 5 | 1 | 0 | Moderate | 0 | 0 | 2 | 5 | Low |
| 1 | 5 | 0 | 1 | Moderate | 0 | 0 | 1 | 6 | Low |
| 1 | 4 | 2 | 0 | Moderate | 0 | 0 | 0 | 7 | Low |

\*High quality (green) when $0 \leq X \leq 0.33$ ; moderate quality (yellow) when $0.33 < X \leq 0.66$ ; low quality (red) when $0.66 < X \leq 1.00$ .

**Figure S1.**
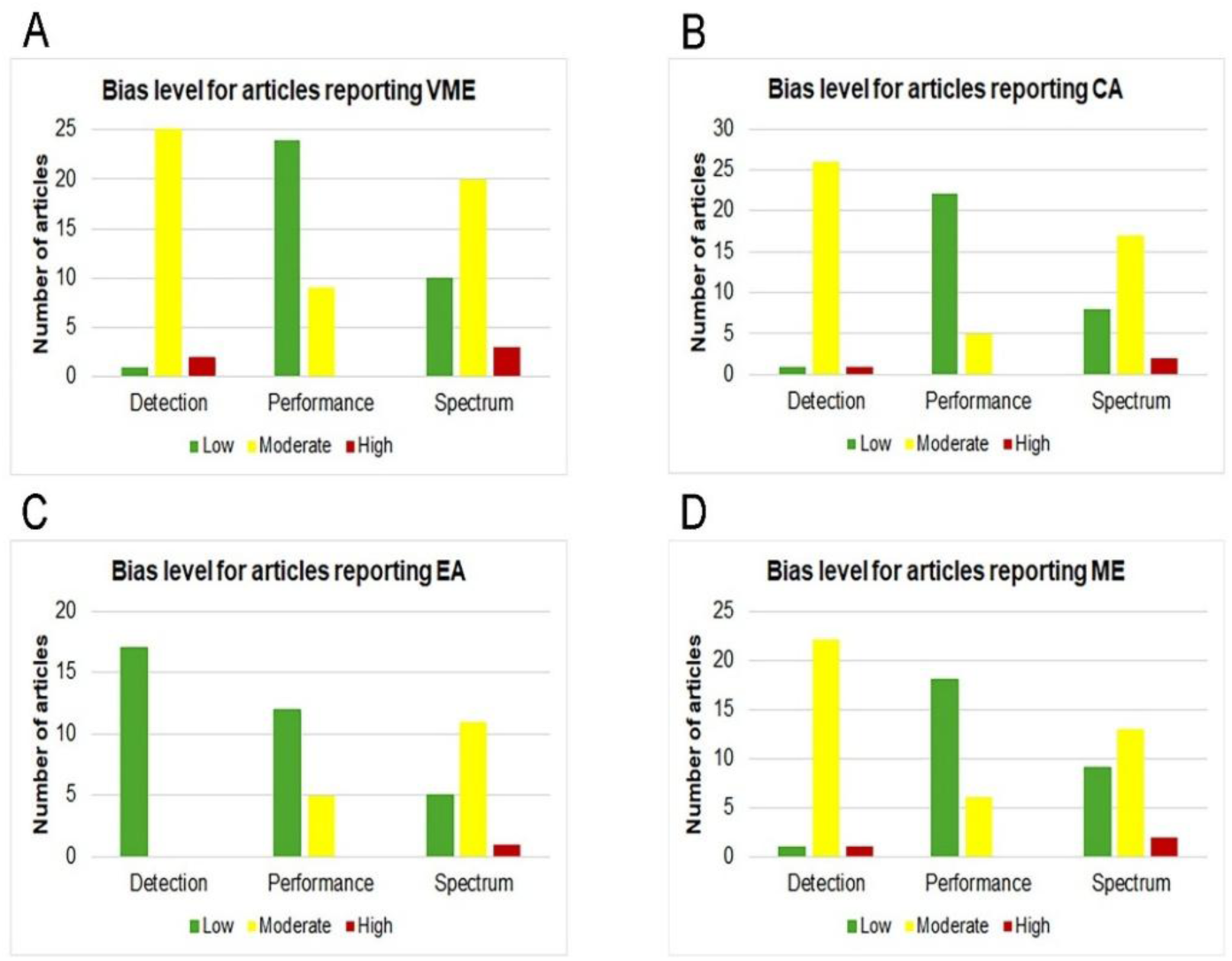
Bias levels for articles within each reporting category (VME, CA, EA, ME). Green, yellow, and red bars indicate low, moderate, and high bias, respectively. See Materials and Methods for details on bias scoring and its integration into overall assessment of study quality (Table 1). **Abbreviations**: EA, essential agreement; CA, categorical agreement; VME, very major error; ME, major error.

**Figure S2.**
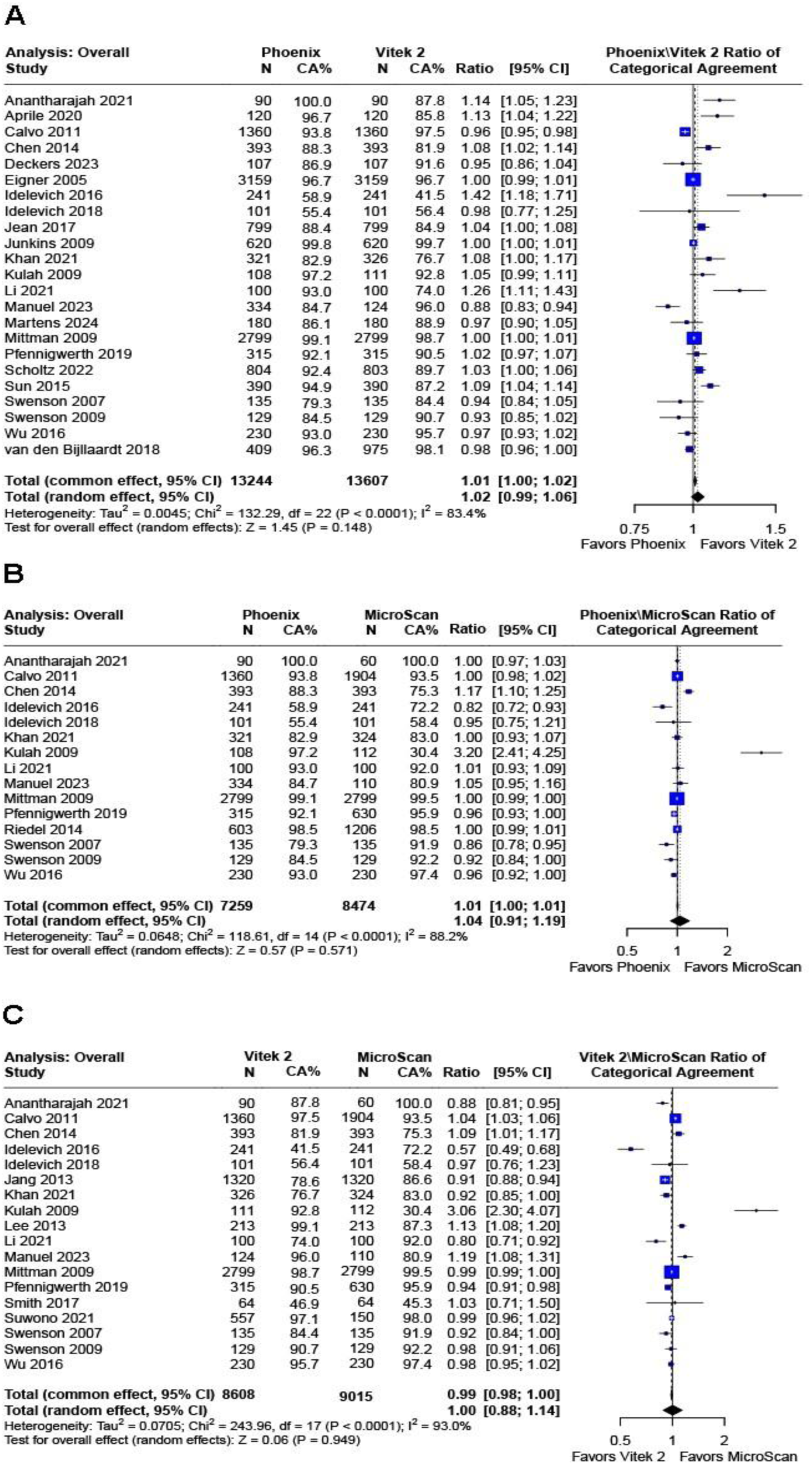
Overall analyses, which included any article reporting categorical agreement values for ≥2 instruments (with a common reference method). Forest plots for Index X / Index Y (X and Y represent Phoenix and/or Vitek 2 and/or MicroScan) categorical agreement ratios following conversion to weighted averages and statistical calculation for common and random effects. The bars spanning from each of the square point estimates represent confidence intervals. The size of the square plots represents the relative size of the group numbers for each included study. p<0.05 was the threshold for statistical significance. (A) (B) and (C) represent comparisons for Phoenix versus Vitek 2, Phoenix versus MicroScan, and Vitek 2 versus MicroScan, respectively. **Abbreviations:** CA, categorical agreement

**Figure S3.**
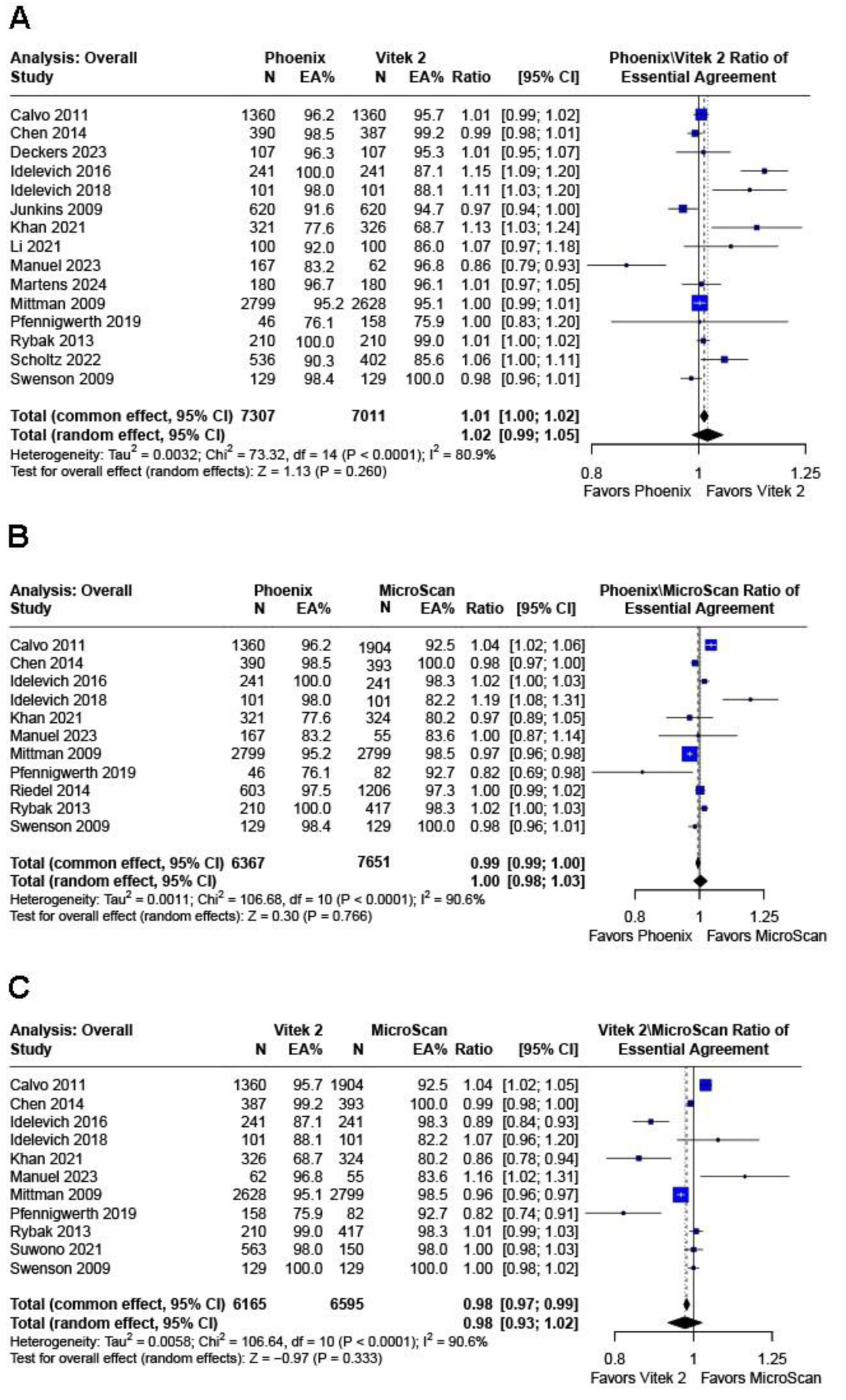
Overall analyses, which included any article reporting essential agreement values for ≥2 instruments (with a common reference method). Forest plots for Index X / Index Y (X and Y represent Phoenix and/or Vitek 2 and/or MicroScan) essential agreement ratios following conversion to weighted averages and statistical calculation for common and random effects. The bars spanning from each of the square point estimates represent confidence intervals. The size of the square plots represents the relative size of the group numbers for each included study. p<0.05 was the threshold for statistical significance. (A) (B) and (C) represent comparisons for Phoenix versus Vitek 2, Phoenix versus MicroScan, and Vitek 2 versus MicroScan, respectively. **Abbreviation:** EA, essential agreement

**Figure S4.**
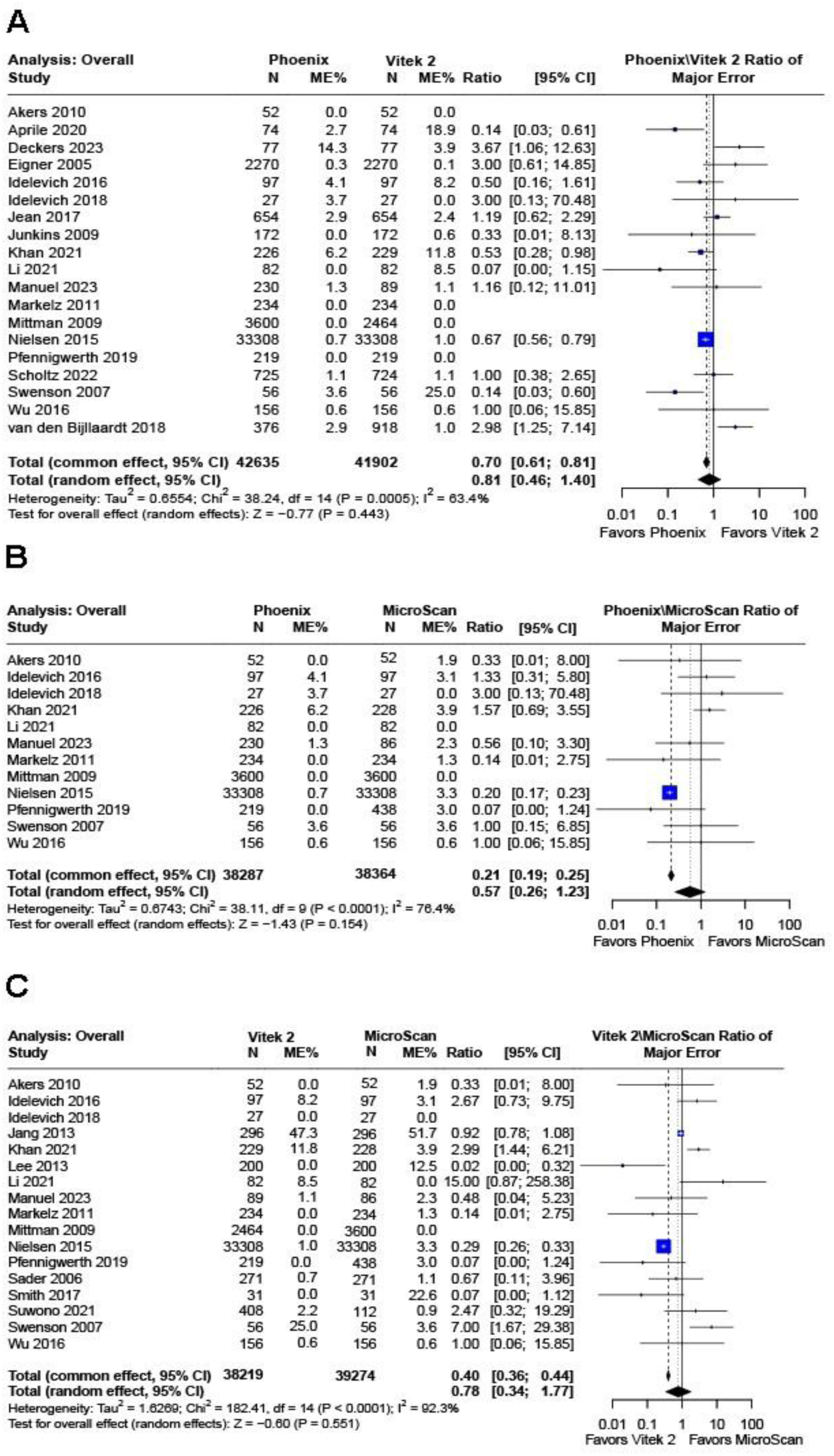
Overall analyses, which included any article reporting major error values for ≥2 instruments (with a common reference method). Forest plots for Index X / Index Y (X and Y represent Phoenix and/or Vitek 2 and/or MicroScan) major error ratios following conversion to weighted averages and statistical calculation for common and random effects. The bars spanning from each of the square point estimates represent confidence intervals. The size of the square plots represents the relative size of the group numbers for each included study. p<0.05 was the threshold for statistical significance. (A) (B) and (C) represent comparisons for Phoenix versus Vitek 2, Phoenix versus MicroScan, and Vitek 2 versus MicroScan, respectively. **Abbreviations**: ME, major error

**Figure S5.**
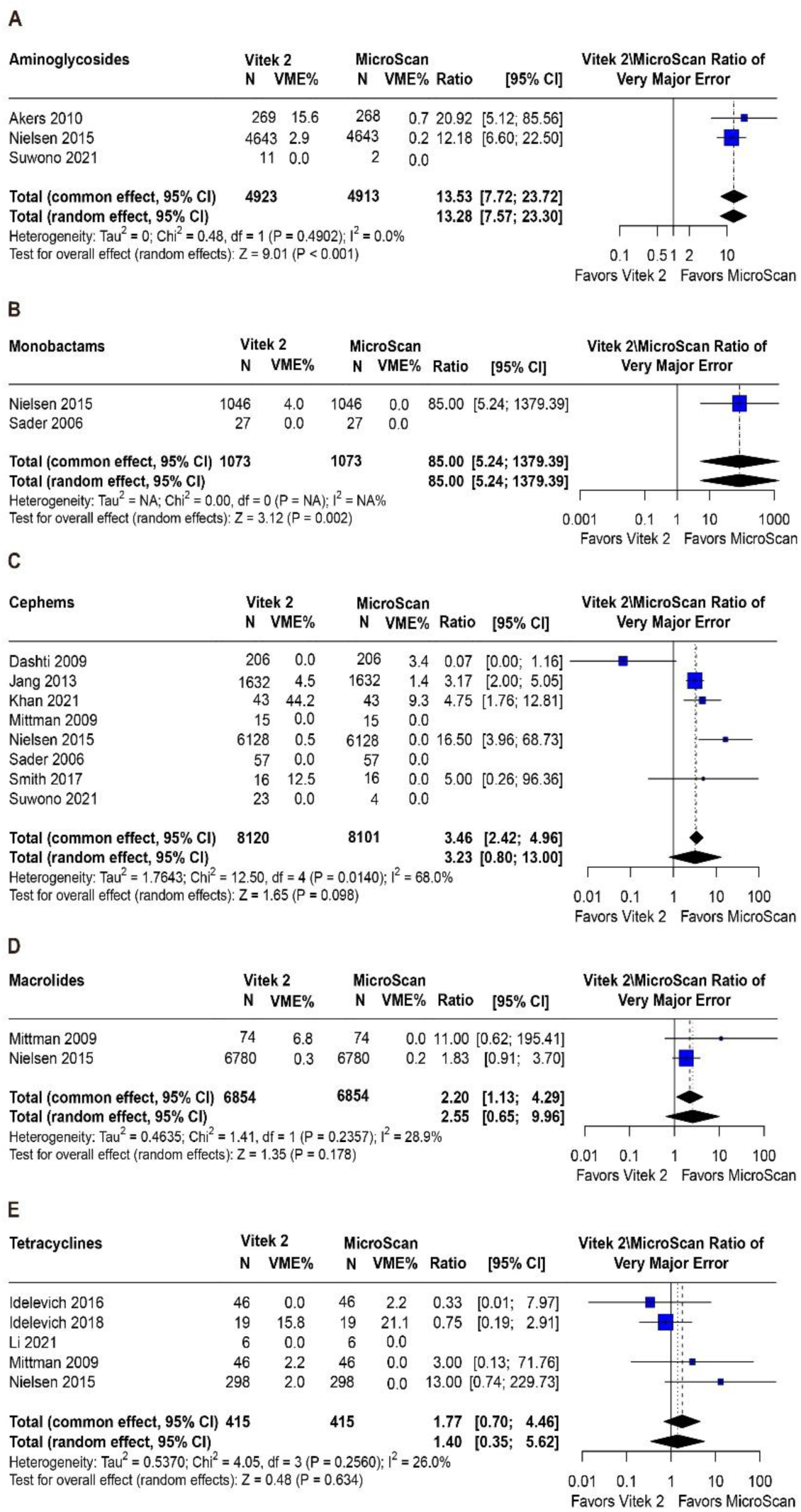
Analysis of five antibiotic drug classes associated with relatively high VME rates for Vitek 2. Forest plots for Vitek 2 - MicroScan associated with very major error ratios following conversion to weighted averages and statistical calculation for common and random effects. The bars spanning from each of the square point estimates represent confidence intervals. The size of the square plots represents the relative size of the group numbers for each included study. p<0.05 was the threshold for statistical significance. (A-E) represent Vitek 2 - MicroScan ratios associated with either aminoglycosides, monobactams, cephems, macrolides, or tetracyclines (A-E, respectively) —the five worst performing drug classes for Vitek 2. **Abbreviations**: VME, very major error

**Figure S6.**
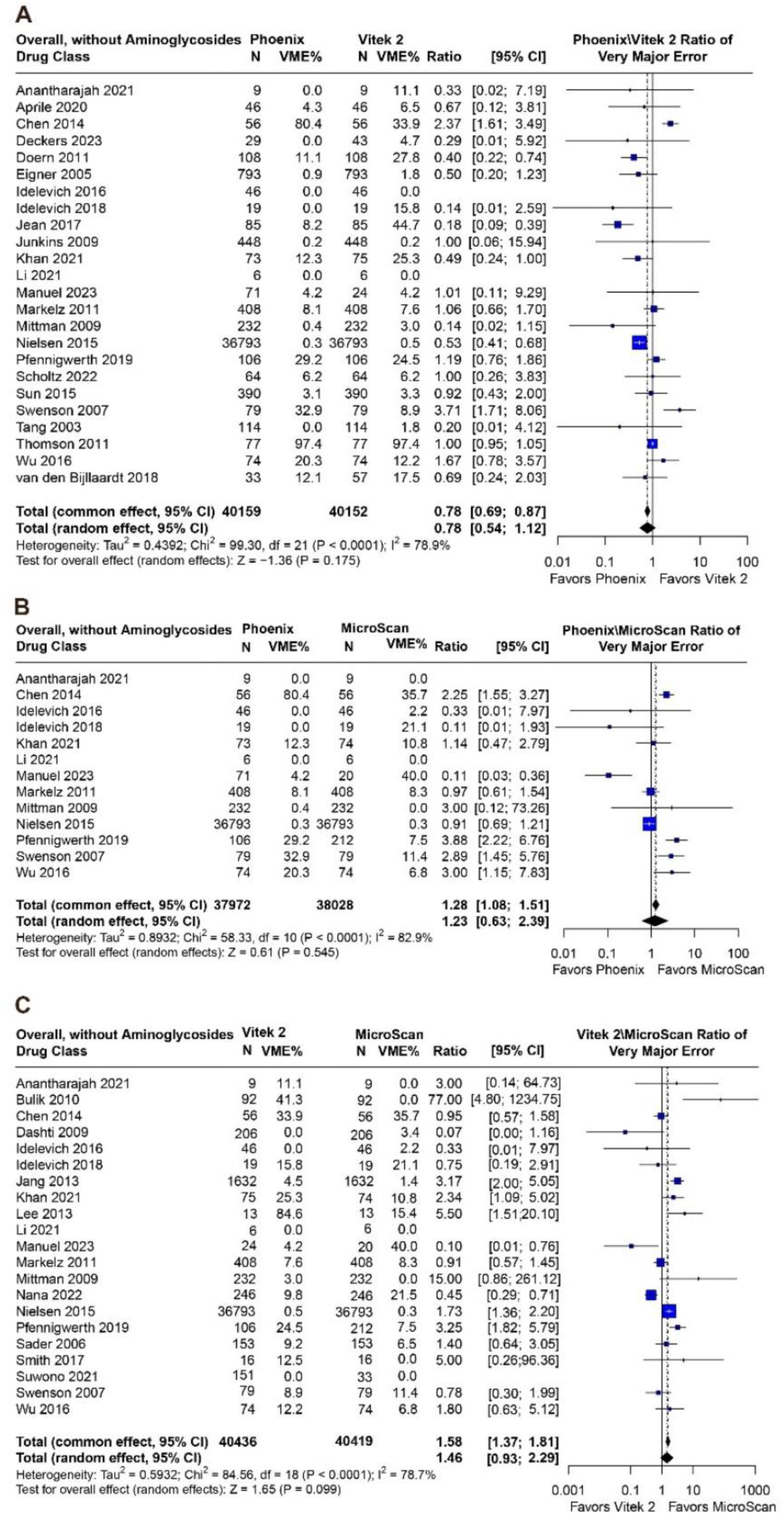
Removal of aminoglycoside drug class from the analysis. Forest plots for VME ratios following conversion to weighted averages and statistical calculation for common random effects. Removal of aminoglycoside, the worst performing drug class for Vitek 2, resulted in no significant difference between Vitek 2 and MicroScan for VME rates. **Abbreviation:** VME, very major error

**Figure S7.**
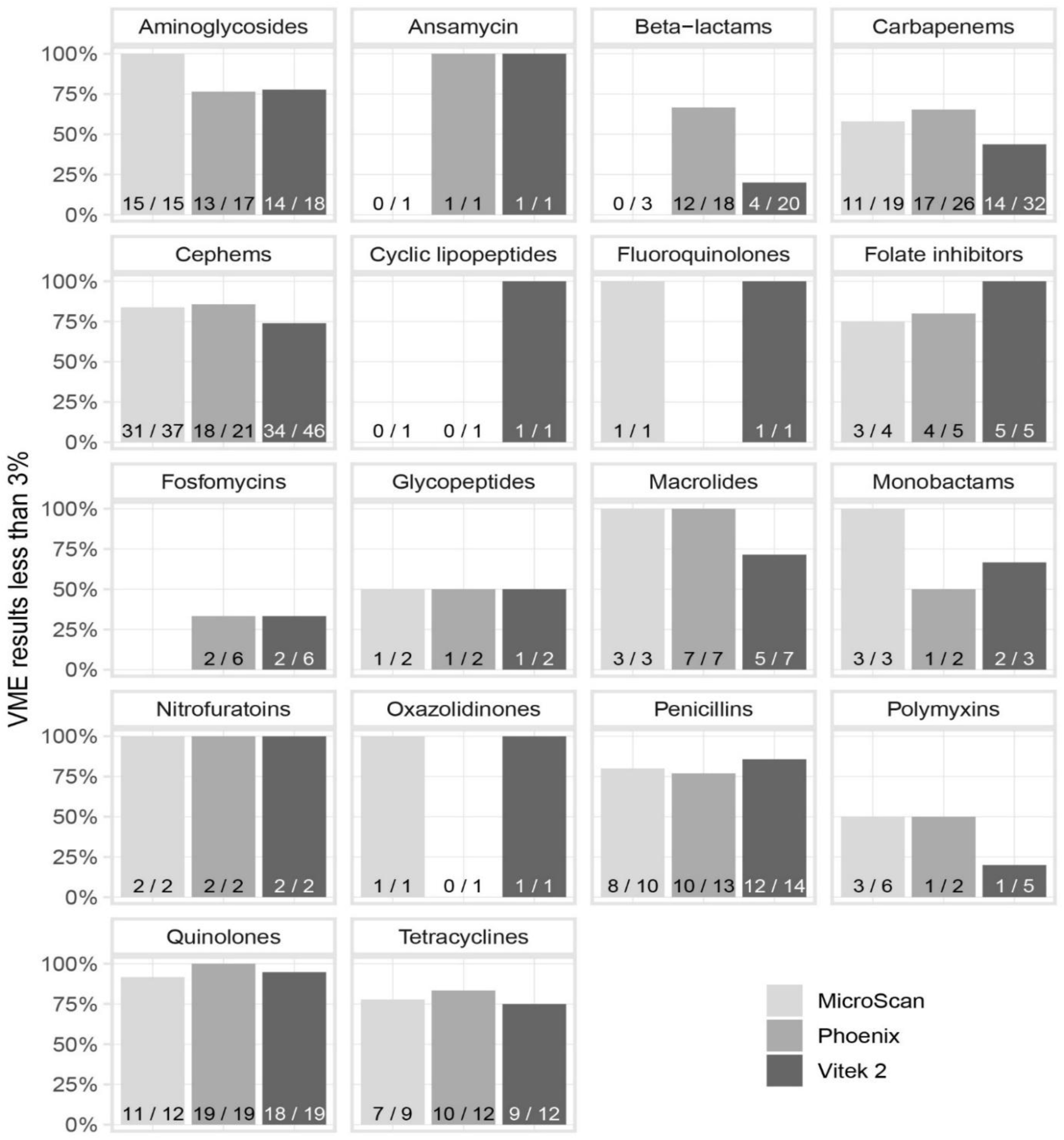
For articles reporting the indicated drug classes, the percentage of those articles having all VME values <3% is plotted (y-axis). Analysis is shown by each of the three AST instruments. For example, of the 18 articles reporting VME values for beta-lactams for Phoenix, 66.7% (12/18) had no values above 3%. **Abbreviation**: VME, very major error

**Figure S8.**
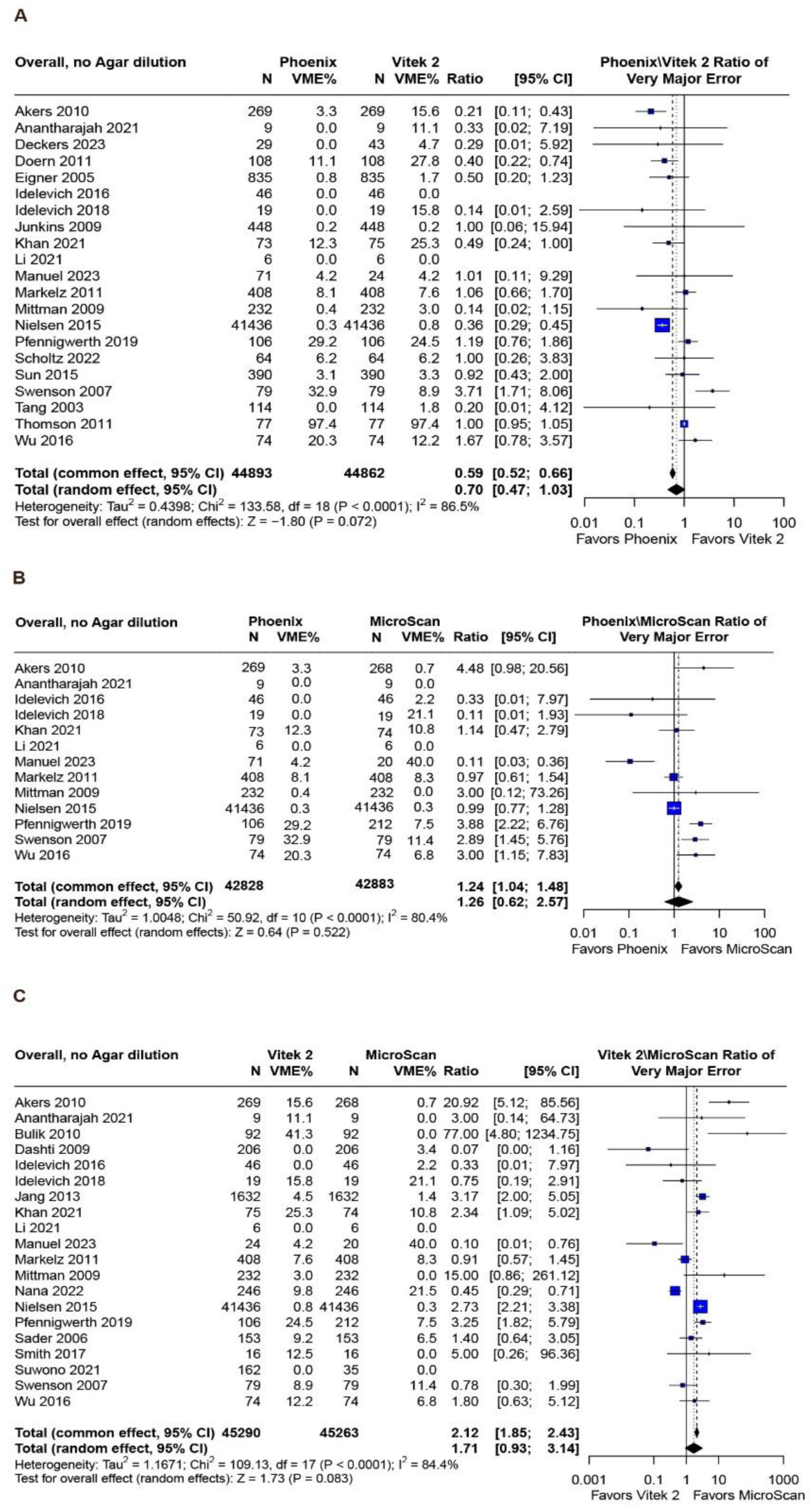
Articles reporting VME values after excluding studies employing agar dilution as the reference method. Forest plots Index X / Index Y (X and Y represent Phoenix and/or Vitek 2 and/or MicroScan) ratios following conversion to weighted averages and statistical calculation for common and random effects. The bars spanning from each of the square point estimates represent confidence intervals. The size of the square plots represents the relative size of the group numbers for each included study. p<0.05 was the threshold for statistical significance. (A) (B) and (C) represent comparisons for Phoenix versus Vitek 2, Phoenix versus MicroScan, and Vitek 2 versus MicroScan, respectively. **Abbreviation:** VME, very major error

**Figure S9.**
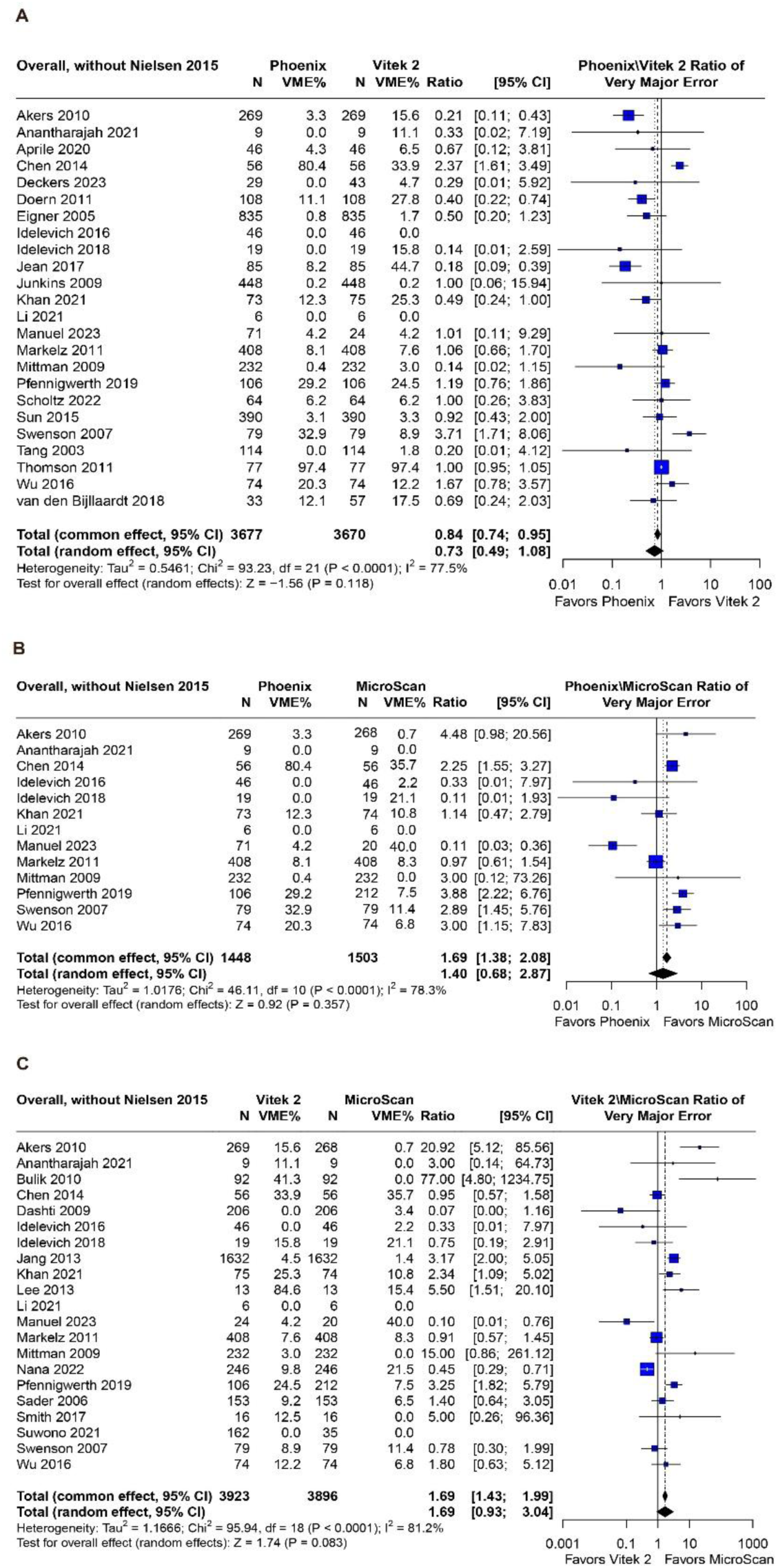
Overall VME (all articles) except for Nielsen 2015. Forest plots Index X / Index Y (X and Y represent Phoenix and/or Vitek 2 and/or MicroScan) ratios following conversion to weighted averages and statistical calculation for common and random effects. The bars spanning from each of the square point estimate represent confidence intervals. The size of the square plots represents the relative size of the group numbers for each included study. p<0.05 was the threshold for statistical significance. (A) (B) and (C) represent comparisons for Phoenix versus Vitek 2, Phoenix versus MicroScan, and Vitek 2 versus MicroScan, respectively. **Abbreviation:** VME, very major error

## REFERENCES

1. WHO. Antimicrobial resistance. https://www.who.int/news-room/fact-sheets/detail/antimicrobial-resistance. Accessed 07/28/2025.

2. Murray CJL, Ikuta KS, Sharara F, et al. Global burden of bacterial antimicrobial resistance in 2019: a systematic analysis. The Lancet. 2022;399(10325):629–655. doi:10.1016/S0140-6736(21)02724-0

3. Prevention USCfDCa. National Antimicrobial Resistance Monitoring System (NARMS). https://www.cdc.gov/narms/about/index.html. Accessed 07/28/2025.

4. WHO. Report signals increasing resistance to antibiotics in bacterial infections in humans and need for better data. https://www.who.int/news/item/09-12-2022-report-signals-increasing-resistance-to-antibiotics-in-bacterial-infections-in-humans-and-need-for-better-data. Accessed 07/28/2025.

5. Fournier PE, Drancourt M, Colson P, Rolain JM, La Scola B, Raoult D. Modern clinical microbiology: new challenges and solutions. Nat Rev Microbiol. Aug 2013;11(8):574–85. doi:10.1038/nrmicro3068

6. Cherie N, Berta DM, Tamir M, et al. Improving laboratory turnaround times in clinical settings: A systematic review of the impact of lean methodology application. PLoS One. 2024;19(10):e0312033. doi:10.1371/journal.pone.0312033

7. Culbreath K, Piwonka H, Korver J, Noorbakhsh M. Benefits Derived from Full Laboratory Automation in Microbiology: a Tale of Four Laboratories. J Clin Microbiol. Feb 18 2021;59(3)doi:10.1128/jcm.01969-20

8. Zhou M, Wang Y, Liu C, et al. Comparison of five commonly used automated susceptibility testing methods for accuracy in the China Antimicrobial Resistance Surveillance System (CARSS) hospitals. Infect Drug Resist. 2018;11:1347–1358. doi:10.2147/idr.S166790

9. Vitek 2 User Manual. bioMérieux. Durham, NC; 2022.

10. Phoenix User Manual. Becton, Dickinson and Company. Sparks, MD; 2024.

11. DxM MicroScan WalkAway User Manual. Beckman Coulter, Inc. Brea, CA; 2021.

12. Gatsonis C, Paliwal P. Meta-analysis of diagnostic and screening test accuracy evaluations: methodologic primer. AJR Am J Roentgenol. Aug 2006;187(2):271–81. doi:10.2214/ajr.06.0226

13. Leeflang MMG. Systematic reviews and meta-analyses of diagnostic test accuracy. 10.1111/1469-0691.12474. Clinical Microbiology and Infection. 2014/02/01 2014;20(2):105–113. doi:https://doi.org/10.1111/1469-0691.12474

14. Page MJ, McKenzie JE, Bossuyt PM, et al. The PRISMA 2020 statement: an updated guideline for reporting systematic reviews. MetaArXiv. September 14 2020;10.31222/osf.io/v7gm2

15. Booth A, Clarke M, Ghersi D, Moher D, Petticrew M, Stewart L. An international registry of systematic-review protocols. The Lancet. 2011;377(9760):108–109. doi:10.1016/s0140-6736(10)60903-8

16. Wells GA, Shea B, O’Connell D, et al. The Newcastle-Ottawa Scale (NOS) for Assessing the Quality of Nonrandomised Studies in Meta-Analysis. http://www.ohri.ca/programs/clinical_epidemiology/oxford.asp.

17. Schunemann H, Brozek J, Guyatt G, Oxman A. GRADE Handbook for Grading Quality of Evidence and Strength of Recommendations. https://gdt.gradepro.org/app/handbook/handbook.html.

18. delta D. R: A language and environment for statistical computing. R Foundation for Statistical computing, Vienna, Austria. URL https://www.R-project.org/. 2020;

19. Balduzzi S, Rücker G, Schwarzer G. How to perform a meta-analysis with R: a practical tutorial. Evid Based Ment Health. Nov 2019;22(4):153–160. doi:10.1136/ebmental-2019-300117

20. Viechtbauer W. Conducting Meta-Analyses in R with the metafor Package. 2010. 2010–08–05 2010;36(3):48. doi:10.18637/jss.v036.i03

21. Sader HS, Fritsche TR, Jones RN. Accuracy of three automated systems (MicroScan WalkAway, VITEK, and VITEK 2) for susceptibility testing of Pseudomonas aeruginosa against five broad-spectrum beta-lactam agents. J Clin Microbiol. Mar 2006;44(3):1101–4. doi:10.1128/jcm.44.3.1101-1104.2006

22. Nielsen LE, Clifford RJ, Kwak Y, et al. An 11,000-isolate same plate/same day comparison of the 3 most widely used platforms for analyzing multidrug-resistant clinical pathogens. Diagnostic microbiology and infectious disease. 2015;83(2):93–8. doi:10.1016/j.diagmicrobio.2015.05.018

23. CLSI. Breakpoint Implementation Toolkit (BIT). https://clsi.org/resources/breakpoint-implementation-toolkit/ Accessed 08/07/2025.

24. CLSI. Verification of Commercial Microbial Identification and Antimicrobial Susceptibility Testing Systems. 1st ed. CLSI guideline M52. Wayne, PA: Clinical and Laboratory Standards INstitute. 2015;

25. Humphries RM, Ambler J, Mitchell SL, et al. CLSI Methods Development and Standardization Working Group Best Practices for Evaluation of Antimicrobial Susceptibility Tests. J Clin Microbiol. Apr 2018;56(4)doi:10.1128/jcm.01934-17

26. CLSI. Verification of Commercial Microbial Identification and Antimicrobial Susceptibility Testing Systems. 1st ed. CLSI guideline M52. Wayne, PA: Clinical and Laboratory Standards Institute; 2015.

27. Bartoletti M, Antonelli A, Bussini L, et al. Clinical consequences of very major errors with semi-automated testing systems for antimicrobial susceptibility of carbapenem-resistant Enterobacterales. Clin Microbiol Infect. Sep 2022;28(9):1290.e1–1290.e4. doi:10.1016/j.cmi.2022.03.013

28. van Belkum A, Burnham CD, Rossen JWA, Mallard F, Rochas O, Dunne WM, Jr. Innovative and rapid antimicrobial susceptibility testing systems. Nat Rev Microbiol. May 2020;18(5):299–311. doi:10.1038/s41579-020-0327-x

29. Bayot ML, Bragg BN. Antimicrobial Susceptibility Testing. In: StatPearls [Updated 2024 May 17] https://www.ncbi.nlm.nih.gov/sites/books/NBK539714/.

30. Antimicrobial Resistance C. Global burden of bacterial antimicrobial resistance in 2019: a systematic analysis. Lancet. Feb 12 2022;399(10325):629–655. doi:10.1016/S0140-6736(21)02724-0

31. Metlay JP, Waterer GW, Long AC, et al. Diagnosis and Treatment of Adults with Community-acquired Pneumonia. An Official Clinical Practice Guideline of the American Thoracic Society and Infectious Diseases Society of America. Am J Respir Crit Care Med. Oct 1 2019;200(7):e45–e67. doi:10.1164/rccm.201908-1581ST

32. Gupta K, Hooton TM, Naber KG, et al. International clinical practice guidelines for the treatment of acute uncomplicated cystitis and pyelonephritis in women: A 2010 update by the Infectious Diseases Society of America and the European Society for Microbiology and Infectious Diseases. Clinical infectious diseases : an official publication of the Infectious Diseases Society of America. Mar 1 2011;52(5):e103–20. doi:10.1093/cid/ciq257

33. Kalil AC, Metersky ML, Klompas M, et al. Management of Adults With Hospital-acquired and Ventilator-associated Pneumonia: 2016 Clinical Practice Guidelines by the Infectious Diseases Society of America and the American Thoracic Society. Clinical infectious diseases : an official publication of the Infectious Diseases Society of America. Sep 1 2016;63(5):e61–e111. doi:10.1093/cid/ciw353

34. Tamma PD, Hsu AJ. Defining the Role of Novel β-Lactam Agents That Target Carbapenem-Resistant Gram-Negative Organisms. Journal of the Pediatric Infectious Diseases Society. 2019;8(3):251–260. doi:10.1093/jpids/piz002

35. Doern Gary V, Brecher Stephen M. The Clinical Predictive Value (or Lack Thereof) of the Results of In Vitro Antimicrobial Susceptibility Tests. Journal of Clinical Microbiology. 2011/09/01 2011;49(9_Supplement):S11–S14. doi:10.1128/jcm.00580-11

36. Simner PJ, Hindler JA, Bhowmick T, et al. What’s New in Antibiograms? Updating CLSI M39 Guidance with Current Trends. J Clin Microbiol. Oct 19 2022;60(10):e0221021. doi:10.1128/jcm.02210-21

37. Akers KS, Chaney C, Barsoumian A, et al. Aminoglycoside resistance and susceptibility testing errors in Acinetobacter baumannii-calcoaceticus complex. Journal of clinical microbiology. 2010;48(4):1132–8. doi:10.1128/JCM.02006-09

38. Anantharajah A, Glupczynski Y, Hoebeke M, et al. Multicenter study of automated systems for colistin susceptibility testing. European journal of clinical microbiology & infectious diseases : official publication of the European Society of Clinical Microbiology. 2021;40(3):575–579. doi:10.1007/s10096-020-04059-4

39. Aprile A, Scalia G, Stefani S, Mezzatesta ML. In vitro fosfomycin study on concordance of susceptibility testing methods against ESBL and carbapenem-resistant Enterobacteriaceae. J Glob Antimicrob Resist. Dec 2020;23:286–289. doi:10.1016/j.jgar.2020.09.022

40. Bulik CC, Fauntleroy KA, Jenkins SG, et al. Comparison of meropenem MICs and susceptibilities for carbapenemase-producing Klebsiella pneumoniae isolates by various testing methods. J Clin Microbiol. Jul 2010;48(7):2402–6. doi:10.1128/jcm.00267-10

41. Calvo J, Cano ME, Pitart C, et al. Evaluation of three automated systems for susceptibility testing of enterobacteria containing qnrB, qnrS, and/or aac(6’)-Ib-cr. Journal of clinical microbiology. 2011;49(9):3343–5. doi:10.1128/JCM.00563-11

42. Chen SY, Liao CH, Wang JL, et al. Method-specific performance of vancomycin MIC susceptibility tests in predicting mortality of patients with methicillin-resistant Staphylococcus aureus bacteraemia. Journal of Antimicrobial Chemotherapy. 2014;69(1):211–218. doi:10.1093/jac/dkt340

43. Dashti AA, Jadaon MM, Habeeb FM. Can we rely on one laboratory test in detection of extended-spectrum beta-lactamases among Enterobacteriaceae? An evaluation of the Vitek 2 system and comparison with four other detection methods in Kuwait. Journal of clinical pathology. Aug 2009;62(8):739–42. doi:10.1136/jcp.2009.066134

44. Deckers C, Belik F, Denis O, et al. Multicenter interlaboratory study of routine systems for the susceptibility testing of temocillin using a challenge panel of multidrug-resistant strains. European journal of clinical microbiology & infectious diseases : official publication of the European Society of Clinical Microbiology. 2023;42(12):1477–1483. doi:10.1007/s10096-023-04681-y

45. Doern CD, Dunne WM, Jr., Burnham C-AD. Detection of Klebsiella pneumoniae carbapenemase (KPC) production in non-Klebsiella pneumoniae Enterobacteriaceae isolates by use of the Phoenix, Vitek 2, and disk diffusion methods. Journal of clinical microbiology. 2011;49(3):1143–7. doi:10.1128/JCM.02163-10

46. Eigner U, Schmid A, Wild U, Bertsch D, Fahr AM. Analysis of the comparative workflow and performance characteristics of the VITEK 2 and Phoenix systems. J Clin Microbiol. Aug 2005;43(8):3829–34. doi:10.1128/JCM.43.8.3829-3834.2005

47. Idelevich EA, Büsing M, Mischnik A, Kaase M, Bekeredjian-Ding I, Becker K. False non-susceptible results of tigecycline susceptibility testing against Enterobacteriaceae by an automated system: a multicentre study. J Med Microbiol. Aug 2016;65(8):877–881. doi:10.1099/jmm.0.000281

48. Idelevich EA, Freeborn DA, Seifert H, Becker K. Comparison of tigecycline susceptibility testing methods for multidrug-resistant Acinetobacter baumannii. Diagnostic microbiology and infectious disease. 2018;91(4):360–362. doi:10.1016/j.diagmicrobio.2018.03.011

49. Jang W, Park YJ, Park KG, Yu J. Evaluation of MicroScan WalkAway and Vitek 2 for determination of the susceptibility of extended-spectrum β-lactamase-producing Escherichia coli and Klebsiella pneumoniae isolates to cefepime, cefotaxime and ceftazidime. J Antimicrob Chemother. Oct 2013;68(10):2282–5. doi:10.1093/jac/dkt172

50. Jean SS, Liao CH, Sheng WH, Lee WS, Hsueh PR. Comparison of commonly used antimicrobial susceptibility testing methods for evaluating susceptibilities of clinical isolates of Enterobacteriaceae and nonfermentative Gram-negative bacilli to cefoperazone-sulbactam. *Journal of Microbiology*, Immunology and Infection. 2017;50(4):454–463. doi:10.1016/j.jmii.2015.08.024

51. Junkins AD, Lockhart SR, Heilmann KP, et al. BD Phoenix and Vitek 2 detection of mecA-mediated resistance in Staphylococcus aureus with cefoxitin. J Clin Microbiol. 2009;47(9):2879–82. doi:10.1128/JCM.01109-09

52. Khan A, Arias CA, Abbott A, Dien Bard J, Bhatti MM, Humphries RM. Evaluation of the Vitek 2, Phoenix, and MicroScan for Antimicrobial Susceptibility Testing of Stenotrophomonas maltophilia. J Clin Microbiol. Aug 18 2021;59(9):e0065421. doi:10.1128/jcm.00654-21

53. Kulah C, Aktas E, Comert F, Ozlu N, Akyar I, Ankarali H. Detecting imipenem resistance in Acinetobacter baumannii by automated systems (BD Phoenix, Microscan WalkAway, Vitek 2); high error rates with Microscan WalkAway. BMC Infect Dis. Mar 16 2009;9:30. doi:10.1186/1471-2334-9-30

54. Lee SY, Shin JH, Lee K, et al. Comparison of the Vitek 2, MicroScan, and Etest methods with the agar dilution method in assessing colistin susceptibility of bloodstream isolates of Acinetobacter species from a Korean university hospital. J Clin Microbiol. Jun 2013;51(6):1924–6. doi:10.1128/jcm.00427-13

55. Li H, Zhou M, Chen X, et al. Comparative Evaluation of Seven Tigecycline Susceptibility Testing Methods for Carbapenem-Resistant Enterobacteriaceae. Infect Drug Resist. 2021;14:1511–1516. doi:10.2147/IDR.S289499

56. Manuel C, Maynard R, Abbott A, et al. Evaluation of Piperacillin-Tazobactam Testing against Enterobacterales by the Phoenix, MicroScan, and Vitek2 Tests Using Updated Clinical and Laboratory Standards Institute Breakpoints. J Clin Microbiol. 2023;61(2):e0161722. Comment in: J Clin Microbiol. 2023 Nov 21;61(11):e0049723 PMID: 37823655 [https://www.ncbi.nlm.nih.gov/pubmed/37823655] Comment in: J Clin Microbiol. 2023 Nov 21;61(11):e0069223 PMID: 37823659 [https://www.ncbi.nlm.nih.gov/pubmed/37823659]. doi:10.1128/jcm.01617-22

57. Markelz AE, Mende K, Murray CK, et al. Carbapenem susceptibility testing errors using three automated systems, disk diffusion, etest, and broth microdilution and carbapenem resistance genes in isolates of Acinetobacter baumannii-calcoaceticus complex. Antimicrobial Agents and Chemotherapy. 2011;55(10):4707–4711. doi:10.1128/AAC.00112-11

58. Martens S, Cuypers L, Belik F, et al. Multicenter comparison of Etest, Vitek2 and BD Phoenix to broth microdilution for beta-lactam susceptibility testing of Streptococcus pneumonia. Eur J Clin Microbiol Infect Dis. May 27 2024;doi:10.1007/s10096-024-04847-2

59. Mittman SA, Huard RC, Della-Latta P, Whittier S. Comparison of BD phoenix to vitek 2, microscan MICroSTREP, and Etest for antimicrobial susceptibility testing of Streptococcus pneumoniae. Journal of clinical microbiology. 2009;47(11):3557–61. doi:10.1128/JCM.01137-09

60. Nana T, Perovic O, Chibabhai V. Comparison of carbapenem minimum inhibitory concentrations of Oxacillin-48-like Klebsiella pneumoniae by Sensititre, Vitek 2, MicroScan, and Etest. Clin Microbiol Infect. Dec 2022;28(12):1650.e1–1650.e5. doi:10.1016/j.cmi.2022.06.023

61. Pfennigwerth N, Kaminski A, Korte-Berwanger M, et al. Evaluation of six commercial products for colistin susceptibility testing in Enterobacterales. Clin Microbiol Infect. Nov 2019;25(11):1385–1389. doi:10.1016/j.cmi.2019.03.017

62. Riedel S, Neoh KM, Eisinger SW, Dam LM, Tekle T, Carroll KC. Comparison of commercial antimicrobial susceptibility test methods for testing of Staphylococcus aureus and Enterococci against vancomycin, daptomycin, and linezolid. Journal of clinical microbiology. 2014;52(6):2216–22. doi:10.1128/JCM.00957-14

63. Rybak MJ, Vidaillac C, Sader HS, et al. Evaluation of vancomycin susceptibility testing for methicillin-resistant Staphylococcus aureus: comparison of Etest and three automated testing methods. Journal of clinical microbiology. 2013;51(7):2077–81. doi:10.1128/JCM.00448-13

64. Scholtz SL, Faron ML, Buchan BW, Ledeboer NA. Comparison of Methods for Determining the Antibiotic Susceptibility of Aerococcus Species in a Clinical Setting. American Journal of Clinical Pathology. 2022;157(5):781–788. doi:10.1093/ajcp/aqab195

65. Smith KP, Brennan-Krohn T, Weir S, Kirby JE. Improved Accuracy of Cefepime Susceptibility Testing for Extended-Spectrum-Beta-Lactamase-Producing Enterobacteriaceae with an On-Demand Digital Dispensing Method. J Clin Microbiol. Feb 2017;55(2):470–478. doi:10.1128/jcm.02128-16

66. Sun J, Xu Y, Yu Y, Ni Y. Accuracy of in vitro susceptibility tests for carbapenemase-producing Gram-negative bacteria. Journal of medical microbiology. 2015;64(6):620–622. doi:10.1099/jmm.0.000067

67. Suwono B, Hammerl JA, Eckmanns T, et al. Comparison of MICs in Escherichia coli isolates from human health surveillance with MICs obtained for the same isolates by broth microdilution. JAC Antimicrob Resist. Sep 2021;3(3):dlab145. doi:10.1093/jacamr/dlab145

68. Swenson JM, Lonsway D, McAllister S, et al. Detection of mecA-mediated resistance using reference and commercial testing methods in a collection of Staphylococcus aureus expressing borderline oxacillin MICs. Diagnostic microbiology and infectious disease. 2007;58(1):33–9. doi:10.1016/j.diagmicrobio.2006.10.022

69. Swenson JM, Anderson KF, Lonsway DR, et al. Accuracy of commercial and reference susceptibility testing methods for detecting vancomycin-intermediate Staphylococcus aureus. Journal of clinical microbiology. 2009;47(7):2013–7. doi:10.1128/JCM.00221-09

70. Tang P, Low DE, Atkinson S, et al. Investigation of Staphylococcus aureus isolates identified as erythromycin intermediate by the Vitek-1 System: comparison with results obtained with the Vitek-2 and Phoenix systems. J Clin Microbiol. 2003;41(10):4823–5. doi:10.1128/JCM.41.10.4823-4825.2003

71. Thomson KS, Robledo IE, Vazquez GJ, Moland ES. KPC screening by updated BD Phoenix and Vitek 2 automated systems. J Clin Microbiol. 2011;49(9):3386–7. doi:10.1128/JCM.00772-11

72. van den Bijllaardt W, Schijffelen MJ, Bosboom RW, et al. Susceptibility of ESBL Escherichia coli and Klebsiella pneumoniae to fosfomycin in the Netherlands and comparison of several testing methods including Etest, MIC test strip, Vitek2, Phoenix and disc diffusion. The Journal of antimicrobial chemotherapy. 2018;73(9):2380–2387. doi:10.1093/jac/dky214

73. Wu MT, Burnham CAD, Westblade LF, et al. Evaluation of Oxacillin and Cefoxitin Disk and MIC Breakpoints for Prediction of Methicillin Resistance in Human and Veterinary Isolates of Staphylococcus intermedius Group. Journal of clinical microbiology. 2016;54(3):535–42. Comment in: J Clin Microbiol. 2016 Mar;54(3):516-7 PMID: 26763965 [https://www.ncbi.nlm.nih.gov/pubmed/26763965]. doi:10.1128/JCM.02864-15

